# Preparing For the Next Pandemic: Learning Wild Mutational Patterns At Scale For Analyzing Sequence Divergence In Novel Pathogens

**DOI:** 10.1101/2020.07.17.20156364

**Authors:** Jin Li, Timmy Li, Ishanu Chattopadhyay

**Affiliations:** Department of Medicine, University of Chicago; Committee on Genetics, Genomics & Systems Biology, University of Chicago; Committee on Quantitative Methods in Social, Behavioral, and Health Sciences, University of Chicago; Department of Computer Science, University of Chicago, Chicago, IL, USA

## Abstract

As we begin to recover from the COVID-19 pandemic, a key question is if we can avert such disasters in future. Current surveillance protocols generally focus on qualitative impact assessments of viral diversity ^1^. These efforts are primarliy aimed at ecosystem and human impact monitoring, and do not help to precisely quantify emergence. Currently, the similarity of biological strains is measured by the edit distance or the number of mutations that separate their genomic sequences ^2–6^, e.g. the number of mutations that make an avian flu strain human-adapted. However, ignoring the odds of those mutations in the wild keeps us blind to the true jump risk, and gives us little indication of which strains are more risky. In this study, we develop a more meaningful metric for comparison of genomic sequences. Our metric, the q-distance, precisely quantifies the probability of spontaneous jump by random chance. Learning from patterns of mutations from large sequence databases, the q-distance adapts to the specific organism, the background population, and realistic selection pressures; demonstrably improving inference of ancestral relationships and future trajectories. As important application, we show that the q-distance predicts future strains for seasonal Influenza, outperforming World Health Organization (WHO) recommended flu-shot composition almost consistently over two decades. Such performance is demonstrated separately for Northern and Southern hemisphere for different subtypes, and key capsidic proteins. Additionally, we investigate the SARS-CoV-2 origin problem, and precisely quantify the likelihood of different animal species that hosted an immediate progenitor, producing a list of related species of bats that have a quantifiably high likelihood of being the source. Additionally, we identify specific rodents with a credible likelihood of hosting a SARS-CoV-2 ancestor. Combining machine learning and large deviation theory, the analysis reported here may open the door to actionable predictions of future pandemics.

## Introduction

With estimated mortality rates significantly higher compared to that of the seasonal flu, the current COVID-19 pandemic is one of the most devastating disasters of the last 100 years. As researchers strive to develop effective therapeutics and vaccine(s) to combat the SARS-CoV-2 virus, a looming question is if we can be better prepared for the next pandemic. Can we preempt emergence of novel pathogens with an actionable timeline to avert such global devastation the next time around? Current surveillance paradigms, while crucial for mapping disease ecosystems, are limited in their ability to address this challenge. Habitat encroachment, climate change, and other ecological factors ^7–9^ unquestionably drive up the odds of zoonotic spill-over. Nevertheless, efforts at tracking and modeling these effects till date have not improved our ability to quantify future risk of emergence of a specific strain from a specific host ^1^. Existence of viral diversity in hosts such as bats or swines or wild ducks, while important, might not transparently map to emergence risk, and does not address the problem at hand.

Here we argue that a key hurdle to making progress in this direction has been the missing ability to quantitatively assess the risk of emergence from strains that circulate in the wild. The state of the art urgently needs the tools necessary to numerically compute the likelihood of a biological sequence replicating in the wild to spontanously give rise to another by random chance. Indeed, currently the similarity between two genomic sequences is measured by how many mutations it takes to change one to the other. Such a measure does not tell us anything about the true jump-risk. In reality, the odds of one sequence mutating to another is a function of not just how many mutations they are apart to begin with, but also how specific mutations incrementally affect fitness. Without taking into account the constraints arising from the need to conserve function, assessing the jump-likelihood is open to subjective guesswork. Here, we show that a precise calculation is possible: provided the similarity of the sequences is evaluated via a new biology-aware metric, which we call the q-distance.

As applications of the q-distance, we show that 1) learning from the mutational patterns of key surface proteins Hemaglutinnin (HA) and Neuraminidase (NA) for Influenza A (selected for their known roles in cellular entry and exit ^10^), we can improve forecasts for the future dominant circulating strain under seasonal antigenic drift. We outperform WHO’s recommendations for the flu-shot composition almost consistently over past two decades, measured as the number of mutations that separate the predicted from the dominant circulating strain in the target season. Our recommendations repeatedly end up being closer, illustrating the potential of our approach to correctly predict evolutionary trajectories. And, 2) using coding sequences for the surface spike (S) protein, again selected for its known role in cellular entry ^11^, we investigate the SARS-CoV-2 origin problem. We quantify the likelihood of viral strains collected across disparate host species to give rise to the observed SARS-CoV-2 strains, and offer new insights backed by precise numerical assessments not possible with existing tools.

### Material & Methods

Aiming to validate our metric in the context of viral evolution, we begin by collecting relevant coding sequences pertaining to key genes implicated in cellular entry from two public databases (NCBI and GISAID, See Tab. III for tally of total number of distinct sequences used). In this study we use in excess of 30; 000 distinct sequences for betacoronaviruses and Influenza A, focusing on three genes/proteins. For each organism, we uncover a network of dependencies between individual mutations revealed through subtle variations of the aligned sequences. These dependecies define our organism-specific model referred to as the quasi-species network or the Qnet (see Fig. 1 and 2). And the q-distance, informed by the dependencies modeled by the inferred Qnets, adapts to the specific organism, allelic frequencies, and nucleotide variations in the background population. The role of epistatic effects in phenotypic change is well-recognized ^12^; here we factor in such effects in a numerically precise manner to compute bounds on the likelihood of specific strains giving rise to target variants (See Fig. 1b-c).

**TABLE I.** Out-Performance of Qnet recommendations over WHO for Influenza A vaccine composition

| subtype | gene | hemisphere | Two decades (% Improvement) | One decade (% Improvement) |
| --- | --- | --- | --- | --- |
| H1N1 | HA | North | 31.75 | 81.32 |
| H1N1 | HA | South | 33.71 | 72.04 |
| H1N1 | HA | avg | 32.73 | 76.68 |
| H3N2 | HA | North | 39.39 | 41.38 |
| H3N2 | HA | South | 31.00 | 28.81 |
| H3N2 | HA | avg | 35.20 | 35.10 |
| H1N1 | NA | North | 22.09 | 60.00 |
| H1N1 | NA | South | 10.81 | 50.79 |
| H1N1 | NA | avg | 16.45 | 55.40 |
| H3N2 | NA | North | 28.38 | 45.95 |
| H3N2 | NA | South | 24.69 | 47.73 |
| H3N2 | NA | avg | 26.53 | 46.84 |

**Fig 1.**
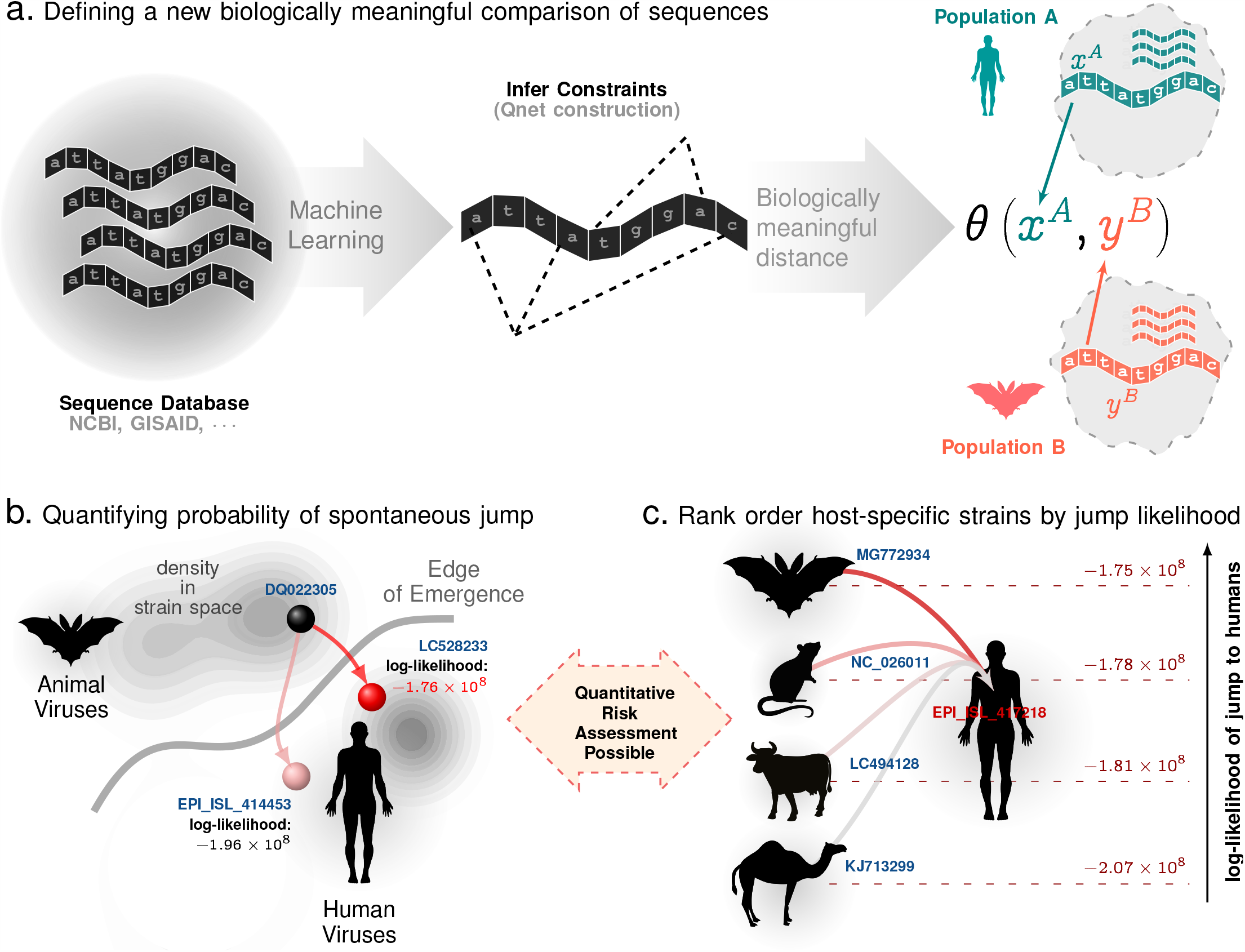
Key insights: Ability to Quantify Risk and Rank-order Strains. Panel **a**. Using sequence variations observed in large databases, we distill evolutionary constraints on a genomic sequence to induce a biology-aware metric for comparing subtle differences in mutating sequences. This metric (q-distance) adjusts to specific organisms, background populations and selection pressures, and reflects the true likelihood of a spontaneous jump from one sequence to the other. We can use this sequence level metric to compute distances between a sequence and a population, and two populations. **Panels b and c** illustrates that we can calculate bounds on the exact likelihood of a spontaneous jump between strains (panel b) and rank-order strains observed in a diverse set of hosts to accurately model future emergence risk (panel c).

**Fig 2.**
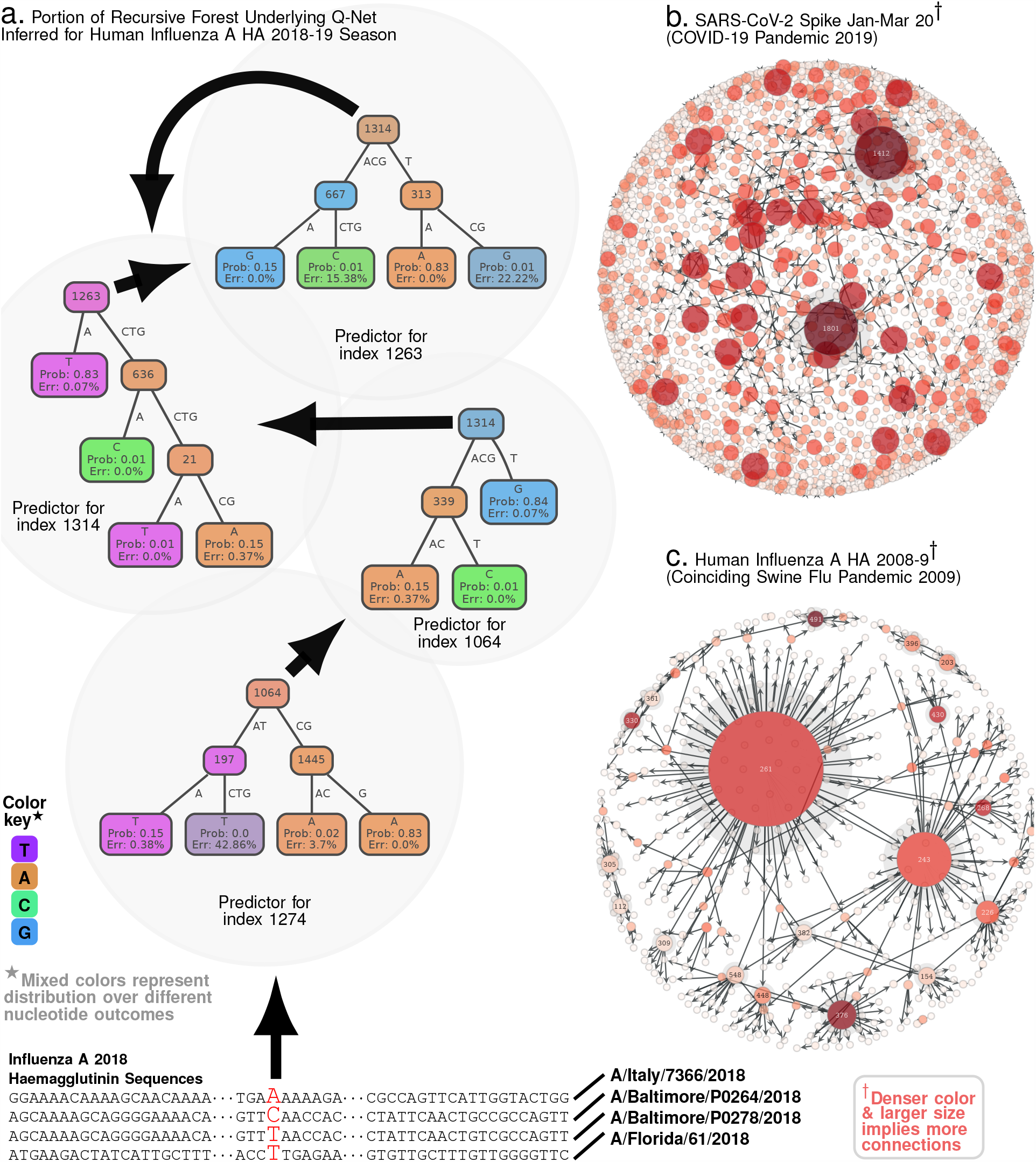
Qnet Computation Scheme. Panel **a**. As an example, beginning with aligned sequences, we calculate a conditional inference tree for index 1274, which involves indices 1064, 1445, 197 as predictive features. These features are automatically selected by the algorithm, as being maximally predictive of the base at 1274. Then, we compute predictors for each of these predictive indices, e:g: we show the inference tree computed for index 1064, which involves index 1314 and 339 as features. Continuing, we find that the predictor for 1314 involves indices 1263, 636 and 21, and that for 1263 involves 1314, 667 and 313. Note that recursive dependencies arise automatically: the predictor for 1263 depends on 1314, and that for 1314 depends on 1263. **Panels b-c** show Qnet dependency graphs for SARS-CoV-2 spike protein and Influenza A HA respectively, illustrating the distinct patterns of mutational constraints inferred. Both HA in Influenza A and the spike protein in SARS-CoV-2 are implicated in viral entry into host cells, and crucial for host specificity of infections. Additionally, the inferred structures underscore the significantly more complex dependencies in SARS-CoV-2 compared to Influenza A.

Clearly in any surveillance effort, we may only observe sequences of high fitness, and only a small subset of viable sequences are ever isolated. A single 10KB observed equence represents a single observation in a 10,000 dimensional space; thus we might never collect enough data points to exhaustively model the set of epistatic dependencies for any realistic genome length. Nevertheless, our results indicate that the scientific community has now accumulated enough sequences for us get meaningful results, at least for some RNA viruses with high mutational rates that reveal enough of the hidden constraints. Admittedly this is but one piece of the puzzle: putting numbers in place of qualitative judgments does not automatically resolve the complex modeling problem of emergence ^13–20^. Notwithstanding the limitations, the ability to quantitatively contrast sequence similarity addresses key aspects of this problem, allowing us to carry out precise comparisons not possible before.

To design our metric, we employ a suite of customized machine learning algorithms to infer the Qnet from aligned genomic sequences sampled from the similar populations, *e.g*. HA from Human Influenza A in year 2008, or the spike protein from all bat betacoronaviruses. The Qnet predicts the nucleotide distribution over the base alphabet (the four nucleic acid bases ATGC) at any specific index, conditioned on the nucleotides making up the rest of the sequence of the gene or genome fragment under consideration. We define the q-distance (See Eq. (3) in Methods) as the square-root of the Jensen-Shannon (JS) divergence ^21^ of these conditional distributions from one sequence to another, averaged over the entire sequence. Invoking Sanov’s theorem on large deviations ^21^ (See Methods), we show that the likelihood of spontaneous jump is bounded above and below by a simple exponential function of the q-distance.

The mathematical intuition behind relating the new distance to jump-probability is the same as in the prediction of a biased outcome when we sequentially toss a fair coin. With an overwhelming probability, such an experiment with a fair coin should result in roughly equal number of heads and tails. However, “large deviations” can happen, and the probability of such rare events is quantifiable ^22^ with existing theory. We show here that the likelihood of a spontaneous transition of a genomic sequence to a substantially different variant by random chance may also be similarly bounded, given we have the Qnet as an estimated model of the evolutionary constraints.

How are Qnets constructed? The key idea is surprisingly simple: we learn models for predicting the mutational variations at each index of the genomic sequence using other indices as features. For example, in Fig. 2a, the predictor for index 1274 uses variation at index 1064 as a feature, and the predictor for index 1064 uses index 1314 as a feature, and so on – ultimately uncovering a recursive dependency structure. Collectively, these inter-dependent predictors represent the constraints that shape evolutionary trajectories driven by selection. The inferred dependencies are illustrated in Fig. 2b-c for SARS-CoV-2 S protein and Influenza A HA respectively, showing that these viruses have markedly different dependency networks for proteins that carry out similar functions (Class I fusion protiens ^23^ mediating cellular entry).

Importantly, the q-distance between two sequences may change if we simply change the background populations, and not the sequences themselves (See Table I for examples, where the distance between two specific Influenza A H1N1 Hemaglutinnin sequences vary when we assume they were collected in different years), and sequences might have a large q-distance and a small edit distance, and vice versa (although on average the two distances tend to be positively correlated, see Tab. II). Hence we construct a new Qnet whenever the background populations are expected to be substantially different, *e.g*., we construct separate Qnets for betacoronavirus S protein sequences isolated from bats, rodents, cattle, non-SARS-CoV-2 human betacoronaviruses, and SARS-CoV-2 strains. For tracking drift in Influenza A, we construct a seasonal Qnet for each subtype and protein that we consider. As an important limitation, the q-distance assumes aligned sequences of identical length (although gaps arising from alignment are acceptable and are modeled as missing data). Thus, the Qnet framework is applicable to closely related sequences, and is well-suited to track subtle changes in evolving viral populations.

Before we enumerate our results, we note that the phylogeny based on the q-distance (q-phylogeny) is potentially distinct from the one constructed using the classical distance. And the jump-probability between two strains connected by a path in a q-phylogenetic tree is bounded above and below by simple exponential functions of the path length (See SI Methods). Thus, smaller phylogenetic distances indicate a higher probability of spontaneous transition and vice versa, making the trees much more useful for interpretation of ancestral relationships and charting possible futures. For example, comparing the phylogenies constructed using the q-distance and the classical metric for all betacoronavirus spike protein sequences from the NCBI and GISAID databases (including those for the novel SARS-CoV-2 strains) in Fig 2a-b, we find that the q-distance leads to cleaner phenotypic separation with clues to SARS-CoV-2 origin.

## Results

Our first application aims to predict dominant strains for the seasonal flu epidemic. Periodic adjustment of the Influenza vaccine components is necessary to account for antigenic drift ^24^. The flu shot is annually prepared at least six months in advance, and comprises a cocktail of historical strains determined by the WHO via global surveillance ^25^, hoping to match the circulating strain(s) in the upcomimg flu season. A variety of hard-to-model effects hinders this prediction, and has limited vaccine effectiveness in recent years ^26^.

We hypothesized that since the probability of a jump or deviation exponentially decreases with an increasing q-distance, the centroid of the strain distribution in our metric will drift slowly. If true, the strain selected closest to the “q”-centroid will be a good approximation of next season’s dominant strain. We tested this hypothesis on past two decades of sequence data for Influenza A (H1N1 and H3N2), with promising results: the q-distance based prediction demonstrably outperforms WHO recommendations by reducing the distance between the predicted and the dominant strain (Fig. 3). Here, we identify the dominant strain to be the one that occurs most frequently, computed as the centroid of the strain distribution observed in a given season in the classical sense (no. of mutations). For H1N1 HA the Qnet induced recommendation outperforms the WHO suggestion by > 31% on average over the last 19 years, and > 81% in the last decade in the northern hemisphere. The gains for NA over the same time periods for H1N1 for the north are > 60% and > 22% respectively. For the southern hemisphere, the gains for H1N1 over the last decade are > 72% for HA, and > 50%. The full table of results is given in Tab. I in the Supplementary text. Fig. 3 illustrates the relative gains computed for both subtypes and the two hemispheres (since the flu season occupy distinct time periods and may have different dominant strains in the northern and southern hemispheres ^24^). Additional improvement is possible if we recommend multiple strains every season for the vaccine cocktail (Fig. 3e,f,k,l). The details of the specific strain recommendations made the Qnet approach for two subtypes (H1N1, H3N2), for two genes (HA, NA) and for the northern and the southerm hemispheres over the previous 19 years are enumerated in the Supplementary Tab. V-XIV.

**Fig 3.**
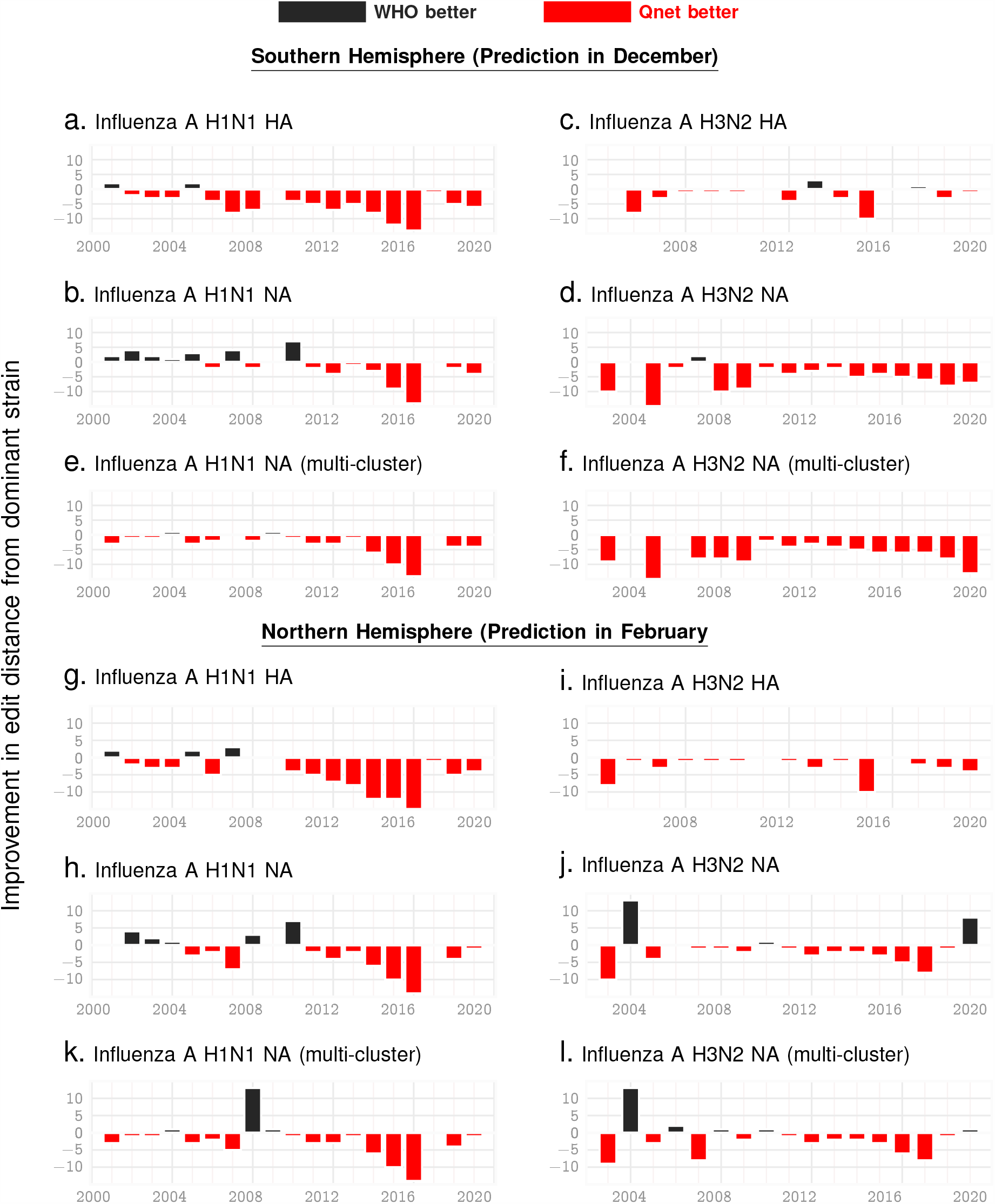
Seasonal Predictions for Influenza. **A**. Relative out-performance of Qnet predictions against WHO recommendations for H1N1 and H3N2 subtypes for the HA and NA coding sequences and over the northern and southern hemispheres. The negative bars (red) indicate the reduced edit distance between the predicted sequence and the actual dominant strain that emerged that year. We see that for the overwhelming majority of seasons, we outperform WHO. Note that the recommendations for the northern hemisphere are given in Februrary, while that for the southern hemisphere are given at teh end of December the previous year, keeping in mind that the flu season in the south begins a few months early. **Panels e**,**f**,**k**,**l** show further possible improvement in NA predictions if we return 3 recommendations instead of one each year.

As our second application of the q-distance approach, we investigate the origin problem of SARS-CoV-2, via quantifying the likelihood of different animal species hosting the immediate progenitor. For any novel pathogen, a plausible history of emergence is generally constructed by estimating similarity of the consensus strain with candidates in suspected animal hosts ^27,28^. However, interpreting a small edit distance as being indicative of a higher chance of a species-jump is problematic, particularly if multiple potential progenitor candidates arise. In contrast, a smaller average q-distance of a novel strain from animal reservoir A vs that from B implies that there is indeed a quantifiably higher probability of a jump from A.

To demonstrate the applicability of this idea, we estimate numerical bounds on the likelihood of the SARS-CoV-2 progenitor arising from specific hosts. Using betacoronavirus sequences from NCBI database corresponding to different animal hosts, we estimate the mean q-distance of SARS-CoV-2 sequences to bats, mouse/rodents, cattle (including camels) and pre-existing human strains including SARS-CoV1, OC43 and HKU1 strains (See Fig. 5a, showing the average log-likelihood of jump from different animal species). We do not a priori restrict our investigation to hosts geographically bound to South East Asia, and demonstrate that this localization arises naturally from our analysis. Our results corroborate the high probability of the progenitor originating from bats as suggested in recent studies ^29,30^ (See Fig. 5a (i), which shows the average lower bound of the log-likelihood of a spontaneous jump from broad host categories to SARS-CoV-2 strains collected upto early March in 2020). In addition, we are also able to identify a ranked list of related bat species with the highest potential of hosting a SARS-CoV-2 progenitor (See Fig 5a (ii), which shows the minimum likelihood of jump to the nearest SARS-CoV-2 strain for the respective host species). Additionally, we find a high likelihood of a close ancestor of SARS-CoV-2 existing in rodents (Fig. 5a).

## Discussion

In this study we formulate a new biology-aware distance between genomic sequences. As a function of this distance, we compute bounds on the explicit probability of a spontaneous jump between nearby variants. We show that quantification of historically qualitative characterizations of ancestral relationaships and future variant calculation improves strain predictions for Influenza A vaccines, and offers new insights into SARS-CoV-2 origin.

High season-to-season genomic variation in the key Influenza capsidic proteins is driven by two opposing influences: 1) the need to conserve function limiting random mutations, and 2) hyper-variability to escape recognition by neutralizing antibodies. Even a single residue change in the surface proteins might dramatically alter recognition characteristics, brought about by unpredictable ^31,32^ changes in local or regional properties such as charge, hydropathy, side chain solvent accessibility ^33–36^. Comparing the Qnet inferred strain (QNT) against the one recommended by the WHO, three important observations come forward: 1) the high likelihood of QNT being closer to the dominant strain (DOM) over the part two decades, and almost consistently over the last decade (See Tab. I and Fig. 3), 2) the residues that only the QNT matches correctly with DOM (while the WHO fails) are largely localized within the receptor binding domain (RBD), with > 57% occurring within the RBD on average (see Fig. 4a for a specific example), and 3) when the WHO strain deviates from the QNT/DOM matched residue, the “correct” residue is often replaced in the WHO recommendation with one that has very different side chain, hydropathy and/or chemical properties (See Fig. 4b-f), suggesting deviations in recognition characteristics. Combined with the fact that we find circulating strains are almost always within a few edits of the DOM (See SI-Fig. 1), these observations suggest that hosts vaccinated with the QNT recommendation is more likely to have season-specific antibodies that are more likely to recognize a larger cross-section of the circulating strains.

**Fig 4.**
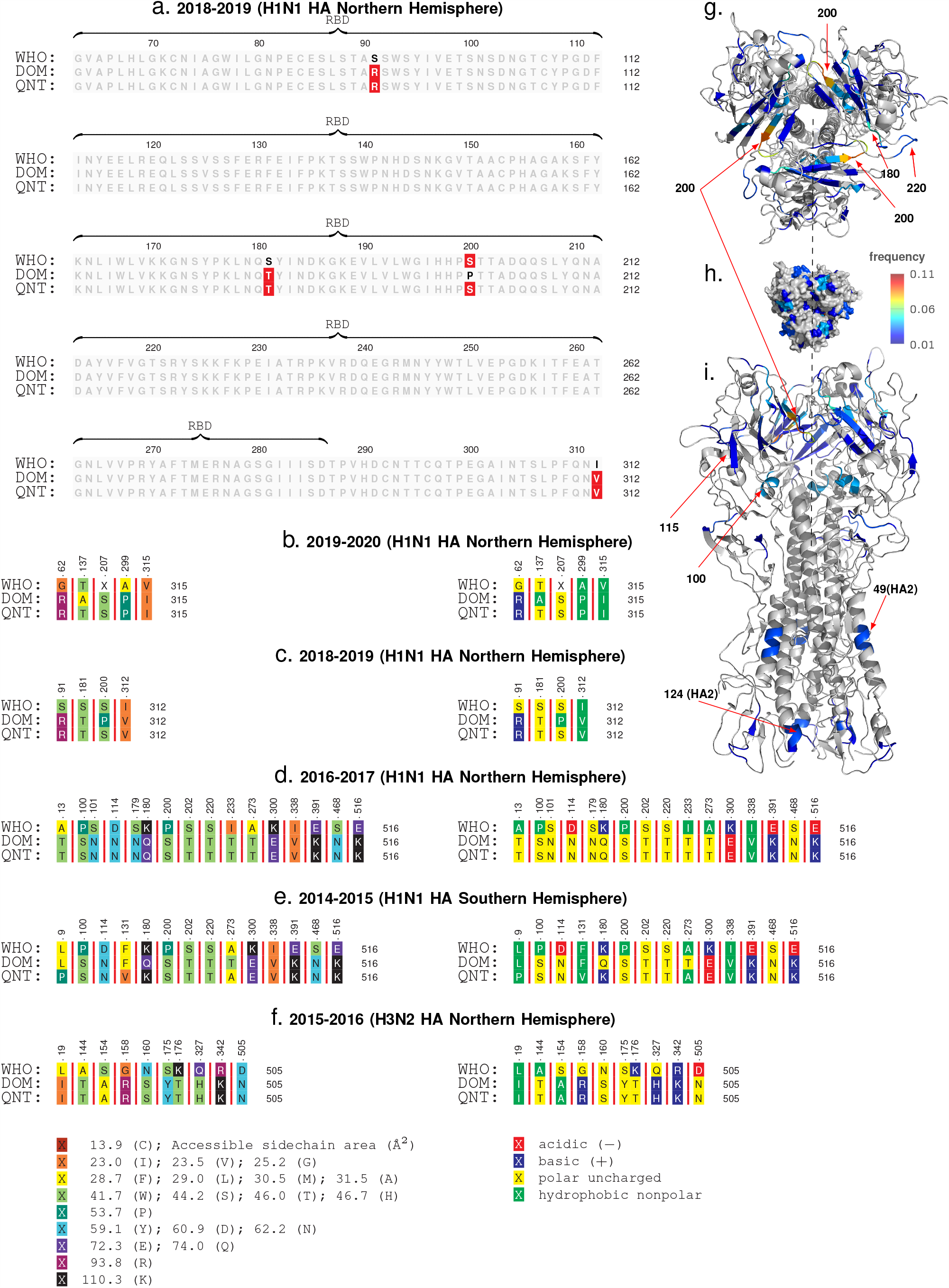
Sequence comparisons. The observed dominant strain, we note that the correct Qnet deviations tend to be within the RBD, both for H1N1 and H3N2 for HA (panel a shows onbe example). Additionally, by comparing the type, side chain area, and the accessible side chain area, we note that the changes often have very different properties (panel b-f). Panels g-i show the localization of the deviationbs in the molecular structure of HA, where we note that the changes are most frequesnt in the HA1 subunit (the globular head), and around residues and structures that have been commonly implicated in receptor binding interactions e.g the 200 loop, the 220 loop and the 180-helix.

Focusing on the average localization of the QNT to WHO deviations in the HA molecular structure, the changes are observed to primarily occur in the HA1 subunit (See Fig. 4g-i, HA0 numbering used, other numbering conversions are given in Supplementary Tab. XVI), with the most frequent deviations occuring around the ≈200 loop, the ≈220 loop, the ≈180 helix, and the ≈100 helix, in addition to some residues in the HA2 subunit (≈49 & ≈124). Unsurprisingly, the residues we find to be most impacted in the HA1 subunit (the globular top of the fusion protein) have been repeatedly implicated in receptor binding interactions ^37–39^. Thus, we are able to fine tune the future recommendation over the state of the art, largely by modifying residue recommendations around the RBD and structures affecting recognition dynamics.

In the context of the origin problem of the 2020 pandemic, we note that literature on SARS-CoV-2 ancestry is still developing, with emerging consensus on horseshoe bats of Chinese origin ^30^ as the potential host of the progenitor sequence. This narrative is primarily driven by observed edit-distance and motif similarities to bat coronavirus (RaTG13, accession MN996532.1) detected in R. affinis from the Yunnan province. However, intriguing questions remain, e.g., the existence of a polybasic furin cleavage site on the spike protein which is absent in RaTG13 and related betacoronaviruses, but do occur in other human coronaviruses including HKU1^30^. Our q-distance analysis (See Fig. 5a) corroborates the progenitor host potential of R. affinis, but we find that a related species R. sinicus is a slightly more probable source. Also, we find several other closely related horseshoe bats including R. ferrumequinum and R. monoceros, and other bats such as T. pachypus, V. superans, and P. abramus are also potential progenitor hosts. In addition, rodents such as R. argentiventer, N. confucianus, and A. agrarius have credible potential as hosting a SARS-CoV-2 ancestor. The top-ranking contenders (excluding humans) ranked by the lower bound of log-likelihood of spontaneous jump to the nearest SARS-CoV-2 strain collected in the relatively early days of the COVID19 pandemic (by early March 2020) is shown in Fig 5a (ii). The role of rodents is further strengthened by noting that SARS-CoV-2 strains and betacoronaviruses from rodents appear in the same clade nested within the clade comprising betacoronaviruses from bats, rodents and SARS-CoV-2 strains (while the rodent strains not being actually closer than those isolated in bats, see Fig. 6)).

**Fig 5.**
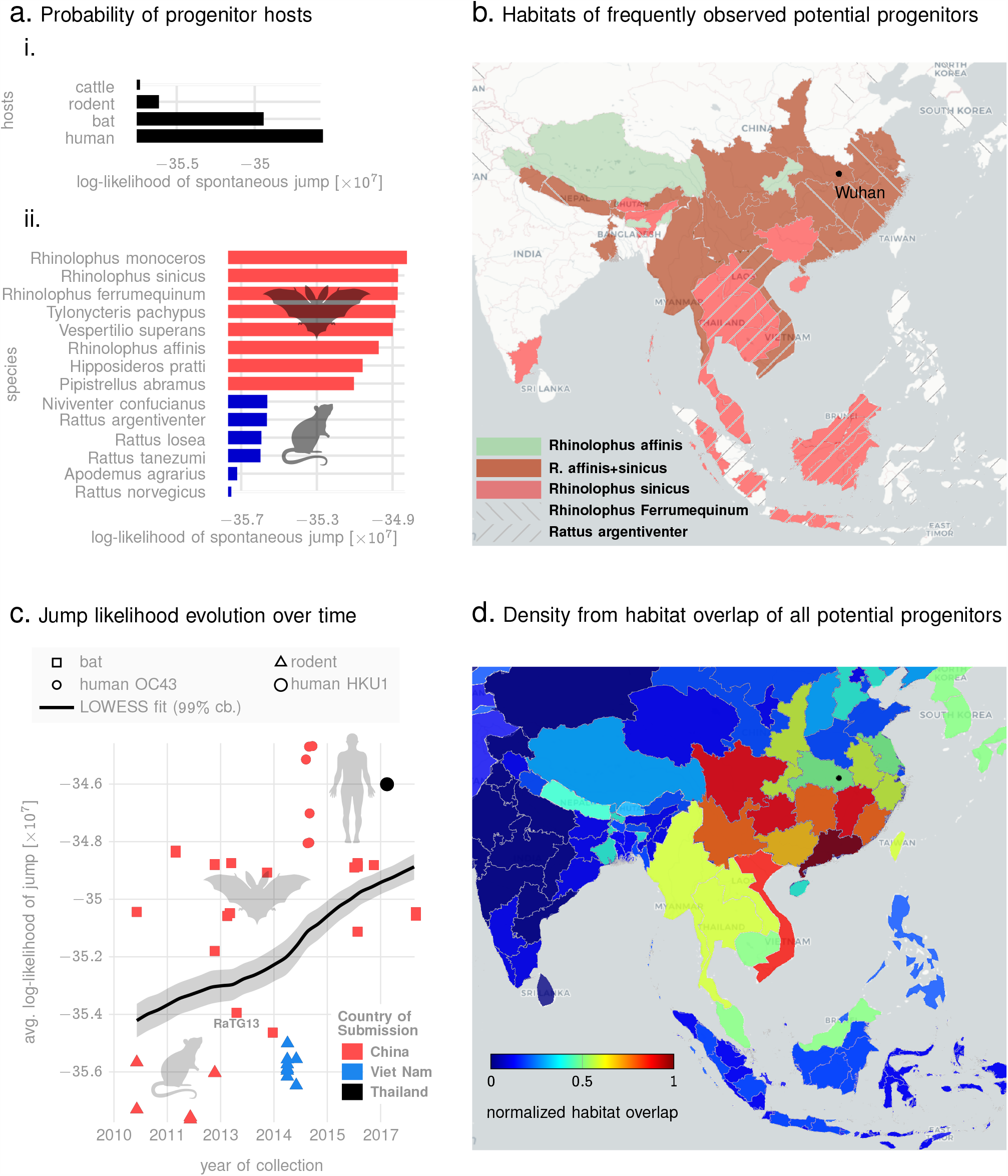
Prediction of animal host for likely progenitors. **Panel a (i)**: average lower bounds on the log-likelihood of jump from different animal hosts to the set of SARS-CoV-2 sequences collected in the early days of the pandemic. **Panel a (ii)**: lower bounds on the log-likelihood of jump from specific species to their respective nearest SARS-CoV-2 neighbors (among sequences collected in the early days of the pandemic). **Panel b** shows the geographic extent of the habitats of the top four most frequently occurring species among the list shown in a(ii). Also, the location of Wuhan, China, ground zero for COVID-19 is shown. **Panel c** plots the lower bound on log-likelihood of various sequences to their nearest neighbors over the time of collection, suggesting a trend of increasing risk over time, and across hosts, as evidenced by a nearly constant gradient LOWESS fit (black line) with 99% confidence bounds. Finally, **panel d** shows the normalized footprint of risk-mediating hosts from overlapping the geographic extents of the habitats of all species from the list in a (ii).

**Fig 6.**
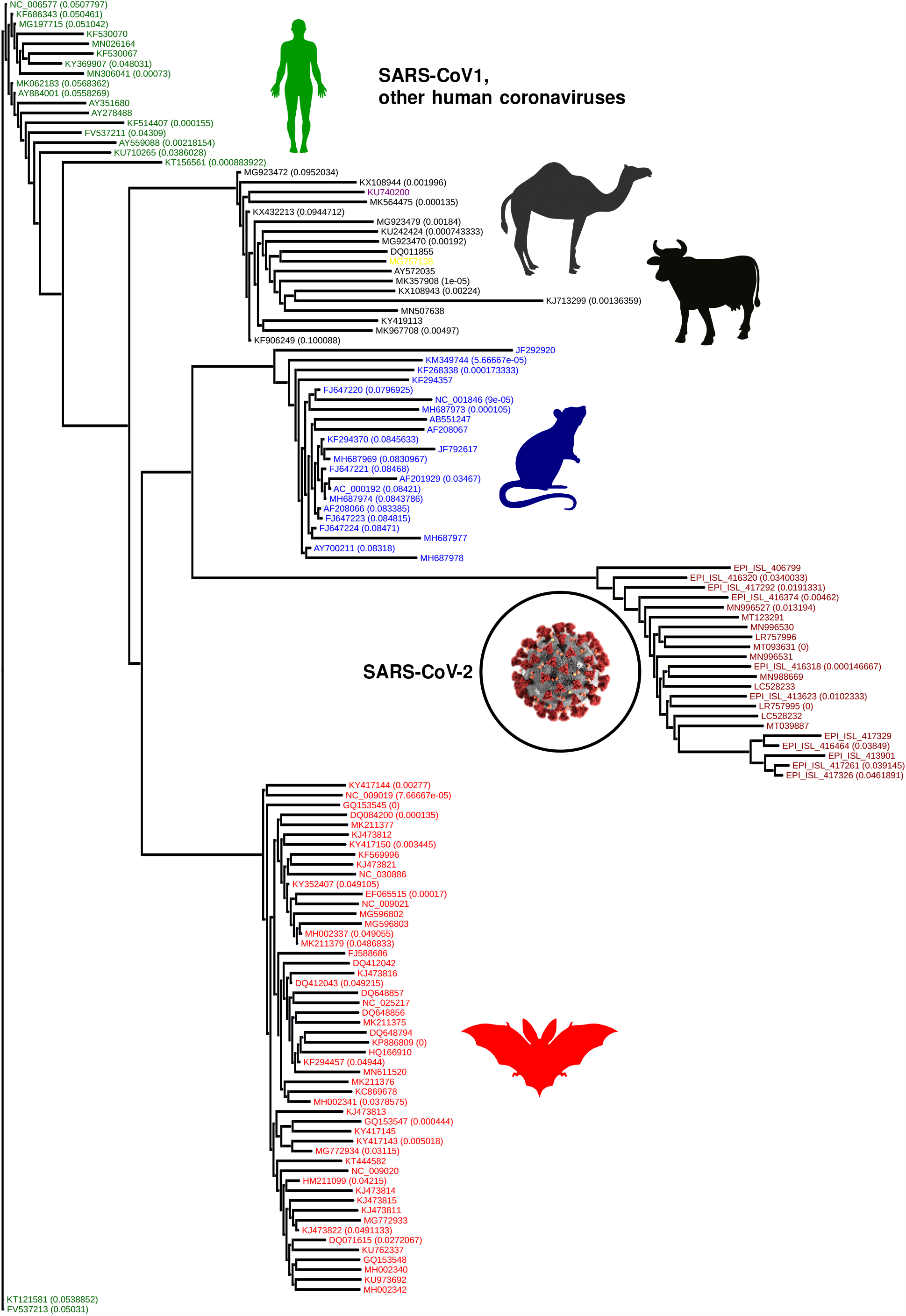
Q-distance induced phylogenetic tree. Importantly, the chronology of SARS-CoV-2 vs existing betacoronaviruses is automatically preserved, and we see the intriguing clade-hierarchy between bat, rodent and SARS-CoV-2 strains. Some branches of the phylogenetic tree is collapsed, and the numbers in bracket list the magnitude of q-distance within which leaves have been collapsed.

In Fig. 5c, we plot the collection times of animal samples against the average lower log-likelihood bound on spontaneous jump to SARS-CoV-2 sequences. We only show sequences that we find to be the top contenders based on their minimum distance from some SARS-CoV-2 sequence collected early in the pandemic (See Table XV). The dependence of the jump probability with collection date suggests risk-progression over time, from at least around 2011. We find that the early risky sequences are exclusively from rodents, and the risk elevates through late 2018, with the majority of the hosts switching from rodents to bats to human coronaviruses (OC43 and HKU1). This progression is further highlighted by a LOWESS regression ^40^ (local polynomial fit to the data points), which shows an almost constant gradient of risk elevation over the past decade. Additionally, overlapping habitats of the top species that pose this risk, we find a normalized habitat distribution (See Fig. 5d) consistent with the presumed ground zero of the outbreak (Wuhan, China).

The quantitative assessments shown in Fig. 5 are impossible in the classical approach, and might suggest that the evolution of SARS-CoV-2 began in rodents, went through bats, and with final maturation in humans. Although we do not provide definitive proof of such a course of events (which realistically might never be found), and these assessments might be impacted by the sparsity of sequences available, the gradual elevation of risk through multiple host species, the overlapping habitats of those species, and the ability to quantify the minimum bounds on jump probability deserve serious consideration.

### Limitations & Conclusion

Calculation of q-distance is currently limited to similar and aligned sequences, e.g. coronoviruses across different hosts, or time frames, or Influenza strains from different subtypes, hosts or seasons. Furthermore, we need a sufficient diversity of observed strains before we can successfully construct the Qnet model; simply having a large number of sequences is not enough, those observations must have sufficient diversity so that the underlying constraints are actually revealed. A multi-variate regression analysis (See SI Methods) indicates that the most important factor for our approach to succeed is indeed the diversity of the sequence dataset, i.e., how many sufficiently distinct sequences have we collected (See Tab. IV). Finally, in the context of strain forecasting, we note that simply reducing the edit distance from the dominant strain is not guaranteed to translate to a better immunological protection. Nevertheless consistent improvement in this metric achieved purely via computational means suggests the possibility of improvement over current practice.

In conclusion, we introduce a data-driven distance metric to track subtle deviations in sequences, and quantify jump risk of risky pathogens. Demonstrated ability of perdicting future flu strains via subtle variations in a limited set of immunologically important residues suggest that the tools developed here could be essential in preempting and actionably mitigating the next pandemic.

## Data Management

Models generated in this study is included as supplementary material, and working software is publicly available at https://pypi.org/project/quasinet/. Accession numbers of all sequences used, and acknowledgement documentation for GISAID sequences in also available as supplementary information.

## Data Availability

Models generated in this study is included as supplementary material, and working software is publicly available at https://pypi.org/project/quasinet/.
Accession numbers of all sequences used, and acknowledgement documentation for GISAID sequences in also available as supplementary information.

https://github.com/zeroknowledgediscovery/quasinet/tree/master/examples

## Acknowledgments

This work is funded in part by the Defense Advanced Research Projects Agency (DARPA) project #FP070943-01-PR. The claims made in this study do not necessarily reflect the position or the policy of the Government, and no official endorsement should be inferred.

## Supplementary Text

### SI Methods

#### Data Source

In this study, we use sequences for the spike (S) protein on betacoronaviruses ^1^, which plays a crucial role in host cellular entry, and the Hemaglutinnin (HA) and Neuraminidase (NA) for Influenza A (for subtypes H1N1 and H3N2), which are key enablers of cellular entry and exit mechanisms respectively ^2^. We use two sequences databases: 1)National Center for Biotechnology Information (NCBI) virus ^3^ and 2) GISAID ^4^ databases. The former is a community portal for viral sequence data, aiming to increase the usability of data archived in various NCBI repositories. GISAID has a somewhat more restricted user agreement, and use of GISAID data in an analysis requires acknowledgment of the contributions of both the Submitting and the Originating laboratories (Corresponding acknowledgment tables are included as Supplementary files). We use a total of 30,204 sequences in our analysis (See Tab. III).

Next, we briefly describe the details of the computational framework.

#### Qnet Framework

In defining the q-distance, we do not assume that the mutational variations at the individual indices of a genomic sequence are independent (See Fig 1a in the main text). Irrespective of whether mutations are truly random ^5^, since only certain combinations of individual mutations are viable, individual mutations across a genomic sequence replicating in the wild appear constrained, which is what is explicitly modeled in our approach. The mathematical form of our metric is not arbitrary; JS divergence is a symmetricised version of the more common KL divergence ^6^ between distributions, and among different possibilities, the q-distance is the simplest metric such that the likelihood of a spontaneous jump (See Eq. (9) in Methods) is provably bounded above and below by simple exponential functions of the q-distance.

Consider a set of random variables *X* = {*X*_*i*_}, with, *i* ∈{1,…,*N*}, each taking value from the respective sets Σ_*i*_. A sample 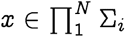 is an ordered *N*-tuple, consisting of a realization of each of the variables *X*_*i*_ with the *i*^th^ entry x_i_ being the realization of random variable *X*_*i*_. We use the notation *x*_−i_ and *x*^*i*,<^ to denote:

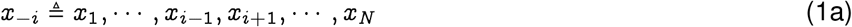

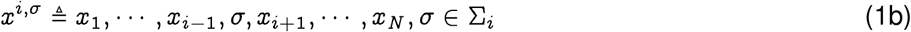

Also, 𝒟(*S*) denotes the set of probability measures on a set *S, e.g*., 𝒟(Σ_*i*_) is the set of distributions on Σ_*i*_.

We note that *X* defines a random field over the index set {1, …, *N*}. Also, to clarify the biological picture, we refer to the sample *x* as an amino acid or nucleotide sequence, identifying the entry at each index with the corresponding protein residue or the nucleotide base pair.

##### Definition 1 (Qnet).

*For a random field X* = {*X*_*i*_} *indexed by i* ∈ {1, …, *N*}, *the Qnet is defined to be the set of predictors* Φ = {Φ_*i*_}, *i.e*., *we have:*

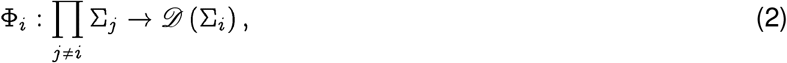

*where for a sequence x*, Φ_*i*_(*x*_−*i*_) *estimates the distribution of X*_*i*_ *on the set Σ*_*i*_.

We use conditional inference trees as models for predictors ^7^, although more general models are possible.

#### Qnet Induced Biology-Aware Distance Between Strains

##### Definition 2 (Pseudo-metric Between Sequences).

*Given two sequences* 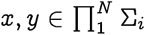, *such that x, y are drawn from the populations P, Q inducing the Qnet* Φ^*P*^, Φ^*Q*^, *respectively, we define a pseudo-metric θ*(*x, y*), *as follows:*

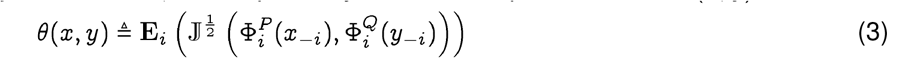

*where* 𝕁 (·, ·) *is the Jensen-Shannon divergence*^8^ *and* ***E***_*i*_ *indicates ation over the indices*.

The square-root in the definition arises naturally from the bounds we are able to prove, and is dictated by the form of Pinsker’s inequality ^6^, making sure that we satisfy the requirement that distances along a path in a constructed phylogeny sum linearly. This allows standard algorithms to be used for phylogeny construction.

Importantly, the *q-distance* defined above is technically a pseudo-metric since distinct sequences can induce the same distributions over each index, and thus evaluate to have a zero distance. This is actually desirable, since we do not want our distance to be sensitive to changes that are not biologically relevant. The intuition is that not all sequence variations brought about by substitutions are equally important or likely. Even with no selection pressure, we might still see random variations at an index if such variations do not affect the replicative fitness. Under that scenario, the corresponding Φ_*i*_ will predict a flat distribution no matter what the input sequence is, thus contributing nothing to the overall distance. And even if two strains *x, y* have the same entry at some index *i*, the remaining residues might induce different distributions Φ_*i*_ based on the remote dependencies, i.e., the entries in *x*_−*i*_, *y*_−*i*_. Also, it matters if the sequences come from two different background populations *P, Q, i.e*., if the induced Qnets Φ^*P*^, Φ^*Q*^ are different. Thus, if we construct Qnets for H1N1 Influenza A separately for the collection years 2008 and 2009, then the same exact sequence collected in the respective years might have a non-zero distance between them, reflecting the fact that the background population the sequences arose from are different, inducing possibly different expected mutational tendencies.

Next, we induce a q-distance between a sequence and a population and between two populations.

##### Definition 3 (Pseudo-metric Between Populations).

*Using the notion of Hausdorff metric between sets:*

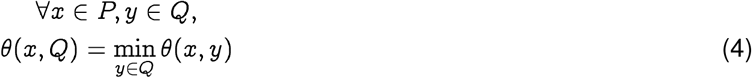

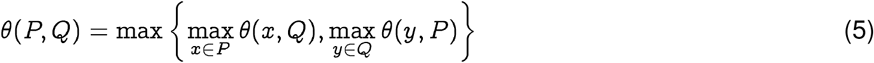

#### In-silico Corroboration of Qnet Constraints

We carry out in-silico experiments to corroborate that the constraints represented within an inferred Qnet are indeed reflective of the biology in play. To that effect, we compare the results of simulated mutational perturbations to sequences from our databases (for which we have already constructed Qnets), and then use NCBI BLAST (https://blast.ncbi.nlm.nih.gov/Blast.cgi) to identify if our perturbed sequences match with existing sequences in the databases (and if so, then where and how many matches they produce). The objective here is to compare such Qnet constrained perturbations against random variations. The results are shown in Fig. 3, where we find that in contrast to random variations, which rapidly diverge the trajectories, the Qnet constraints tend to produce smaller variance in the trajectories, maintain a high degree of match as we extend our trajectories, and produces matches closer in time to the collection time of the initial sequence — suggesting that the Qnet does indeed capture realistic constraints.

#### Significance Test for Population Membership & Progressive Drift in Population Characteristics

For our modeling to be reliable, we need a quantitative test of how well the Qnet represents the data and whether we need to re-calculate the predictors or we have sufficiently many sequences. Here, we formulate an explicit membership test to address this.

##### Definition 4 (Membership Probability of a seuqnce).

*Given a population* P *inducing the Qnet* Φ^*P*^ *and a sequence x, we can compute the membership probability of x:*

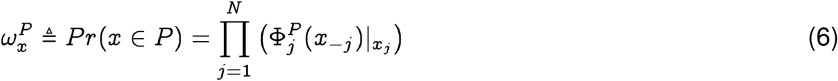

Note that *x*_*j*_ is the *j*^*th*^ entry in *x*, and is thus an element in the set Σ_*j*_. Since we are mostly concerned with the case where Σ_*j*_ is a finite set, 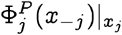 is the entry in the probability mass function corresponding to the element of Σ_*j*_ which appears at the *j*^*th*^ index in sequence *x*.

We can carry out this calculation for a sequence *x* known to be in the population *P* as well, which allows us to define the membership degree 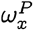.

##### Definition 5 (Membership Degree).

*Let X be a random field representing a population P, ie*.. *X* = *x is a randomly drawn sequence from P*. *Then the membership degree w*^*P*^ *is a function of the random variable X:*

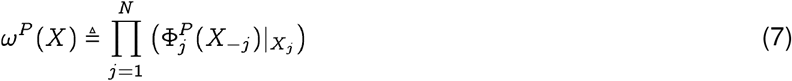

*Note that w*^*P*^ *takes values in the unit interval* [0, 1], *and the probability x is a member of the population P is w*^*P*^(*X* = *x*), *denoted briefly as* 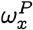 *or w*_*X*_ *if P is clear from context*.

Since *w*^*P*^(*X*) is a random variable, we can now compute sets of sequences that better represent the population P, and ones that are on the fringe. We can also evaluate using a pre-specified significance-level if a particular sequence is not from the population *P*, thus identifying if we need to recompute the predictors Φ, or split the base population. We can set up a hypothesis testing scenario to determine if sequences are indeed from a test population, as follows:

##### Significance Test for the Validity of Inferred Model

Given a population P, inducing a Qnet Φ^*P*^, and a sequence *x*, we assume the null hypothesis is *x* ∉ *P*. We reject the null hypothesis at a pre-specified significance *α*, if

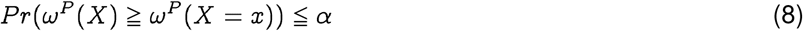

The fraction of newly observed sequences that do not reject the null hypothesis can then be used as an estimate of the species-specific divergence in population characteristics.

The membership degrees for the SARS-CoV-2 sequences in the early days of the pandemic, with respect to our constructed Qnet, is shown in Fig. 4. We find that the distribution of membership degrees is very stable, and almost has no change when we add more sequences (Fig. 4b). In addition, as we collect more sequences, the p-value improves (Fig. 4c), and stabilizes to about 0.02 giving us confidence in the validity of our model.

#### Theoretical Probability Bounds

The Qnet framework allows us to rigorously compute bounds on several quantities of interest, and these bounds are rigorously established in Theorem 1. The fundamental bound is on the probability of a spontaneous change of one strain to another, brought about by chance mutations. While any sequence of mutations is equally likely, the “fitness” of the resultant strain, or the probability that it will even result in a viable strain, is not. Thus the necessity of preserving function dictates that not all random changes are viable, and the probability of observing some trajectories through the sequence space are far greater than others. The Qnet framework allows us to explore this constrained dynamics, as revealed by a sufficiently large set of genomic sequences. With the exponentially exploding number of possibilities in the sequence space, it is computationally intractable to exhaustively model this dynamics. Nevertheless, we can constrain the possibilities using the patterns distilled by the Qnet construction.

We show in Theorem 1 that at a significance level *α*, with a sequence length *N*, the probability of spontaneous jump of sequence *x* from population *P* to sequence *y* in population *Q, Pr*(*x* → *y*), is bounded by:

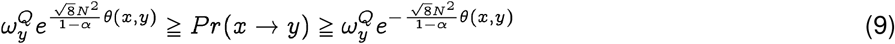

where 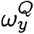 is the membership probability of strain y in the target population.

The ability to estimate the probability of spontaneous jump between sequences in terms of 0 has crucial implications. It allows us to 1) construct a new phylogeny that directly relates the probability of jumps rather than the number of mutations between descendants. 2) simulate realistic trajectories in the sequence space from any given initial strain, and 3) estimate drift in the sequence space by analyzing the statistical characteristics of the diffusion occurring in the strain space.

#### More Fit in the Target Population Makes Jump More Probable

As an immediate consequence of Eq. (9), we can argue that the lower bound of the likelihood of a jump to a target sequence is higher if the final sequence is more fit in the target population. Note that the membership degree by definition quantifies the probability of generating a sequence from our inferred qnet, and since we are far more likely to collect dominant strains when we survey a population, it follows that the membership degree is related to the qualitative notion of fitness.

Conversely, as the fitness of the initial strain (in the neighborhood of 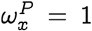) measured by its membership degree falls, the minimum probability of going through a spontaneous jump is higher. We can see this by first noting that for *x* ≠ *y*:

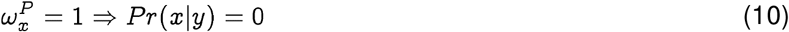

which follows since each term in the product on the right hand side in Eq. (22) is either zero or one if 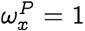, and there is at least one zero since *x* ≠ *y*. To see that the suppression of probability of jump is not simply true if 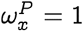 but also in the neighborhood, note that:

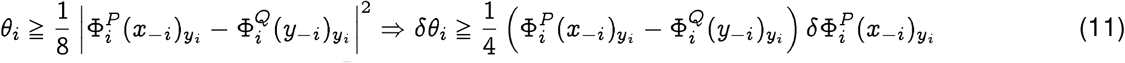

which implies that in the neighborhood of 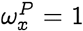, we have:

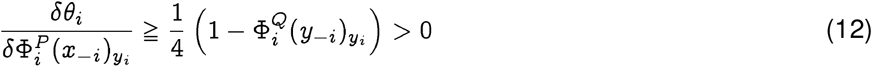

implying that the distance decreases as the membership degree of *x* falls, thus lowering the lower bound on the probability of a spontaneous jump. The argument is not necessarily true if *x* is not in the neighborhood of 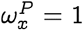 in the first place, and so is of lesser practical interest.

Next, we briefly describe the key applications of the Qnet framework explored in this study, highlighting the predictions made and validations obtained.

#### A Biology Aware Phylogeny

There are more than one computational approach to construct phylogenies, but a majority of these algorithms require a notion of distance between biological sequences, and the edit distance is the one that is most commonly used to construct phylogenies. Using the Qnet induced distance described earlier we can construct phylogenetic trees distinct from those obtained using the classical metric. More importantly, the qnet induced phylogeny is reflective of evolutionary change in a manner that conventional trees are not. As we follow a path in an Q-phylogeny, we can explicitly compute the probability of the changes represented by that path. This probability is bounded above and below by a function of the total path length, i.e., the sum of the q-distances along the path. We can show that for the path *x* = *x*° → … *x*^*k*^ → … *x*^*m*^ = *z*, we have:

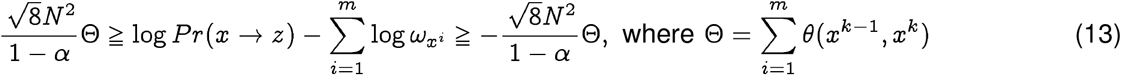

Considering only the lower bound,

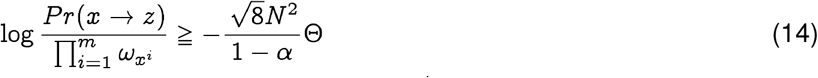

where 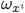 is the membership probability in the base population of the strain *x*^*i*^. Thus, we relate closer phylogenetic distance to explicit probability of spontaneous jump. Note that the definition of the distance function in the Qnet framework allows the summation in Eq. (13), allowing standard tools to construct the phylogenetic tree.

#### Application 1: Predicting Seasonal Strains

Analyzing the distribution of sequences using the q-distance allows us to estimate seasonal drift, which is particularly applicable to Influenza and Influenza-like viruses for which periodic adjustments of vaccine components are necessary to account for antigenic variations.

Our prediction is based on the following intuition: since the probability of spontaneous jump to a strain further away in the q-distance is exponentially lower, the q-centroid of the strain distribution (the centroid computed in the q-distance metric) observed over a season is expected to move slowly, and will be close to the dominant strain in the next season. Thus, we estimate the predicted dominant strain 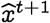 at time *t* + 1, as a function of the observed population at time t as follows:

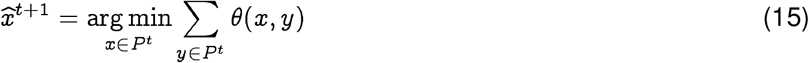

where *P*^*t*^ is the sequence population at time *t*. Here the unit of time is chosen to reflect the appropriate frequency over which vaccine components are re-assessed. In the case of Influenza, this is typically one year. Using this formulation, we test if the predicted strains actually turn out to be closer to the dominant strain in the classical edit distance, when compared against the WHO vaccine recommendation for that season. Our results in Fig. 3 in the main text show that our hypothesis turns out to be correct with few exceptions.

#### Application 2: Identifying Animal Host of Progenitor Sequence

The Qnet based phylogenetic analysis provides a significantly more reliable history of the progenitor strain. In fact, using Eq. (9) we have for the pandemic strain *y* ∈ *H*, and animal strain *x* ∈ *P*:

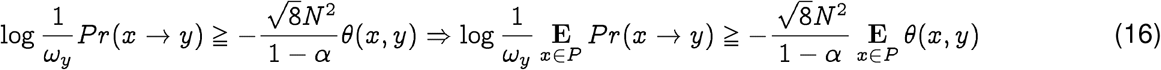

we have constants *C, C*′ such that

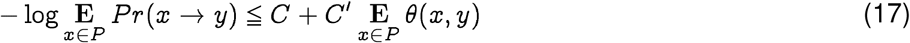

Note that since we always know *N*, we can calculate *C*′ without the knowledge of the pandemic strain *y*. In the case of the SARS-CoV-2 spike protein, at 95% significance, we have:

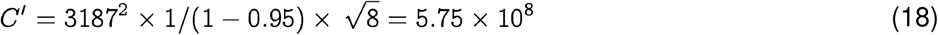

Note that if we have the pandemic strain and are aiming to compare and contrast the likelihood of jump from potential hosts after we already have the emergence event, then we can explicitly calculate *C*. For SARS-CoV-2, this estimate is 4, 805.4 (See Fig. 4), which leads to the following linear relationship between log-likelihood of emergence and the average distance calculated in the Qnet framework:

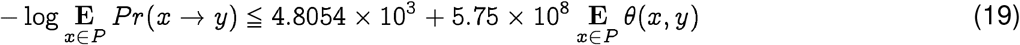

thus providing a quantitative ranking of potential progenitor hosts. It follows from Eq. (19) that for rank-ordering potential hosts, we need to only consider the average distance **E**_*x*∈ *P*_ *θ*(*x, y*). It also follows from the relative magnitudes of the constants in the case of SARS-CoV-2, that we can ignore C and have approximately:

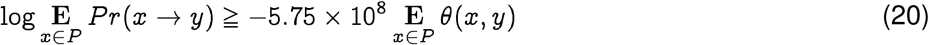

Note that at least in the case of SARS-CoV-2, the fitness term is approximately five orders of magnitude smaller, which implies the jumps probabilities are roughly symmetric. But this is not required to be true in general. At the same time, it is important to note that the probability of jump from strain *x* to strain *y* vs the reverse is actually asymmetric due to the contribution from the population-specific membership degree.

#### Proof of Probability Bounds

**Theorem 1** (Probability Bound). *Given a sequence x of length N that transitions to a strain y* ∈*Q, we have the following bounds at significance level α*.

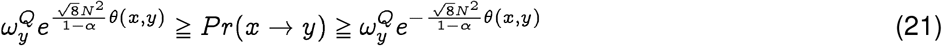

*where* 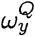 *is the membership probability of strain y in the target population Q*(*See Def. 4*), *and θ*(*x, y*) *is the q-distance between x, y*(*See Def. 2*).

*Proof*. Using Sanov’s theorem^6^ on large deviations, we conclude that the probability of spontaneous jump from strain *x* ∈*P* to strain *y* ∈ *Q*, with the possibility *P* ≠ *Q*, is given by:

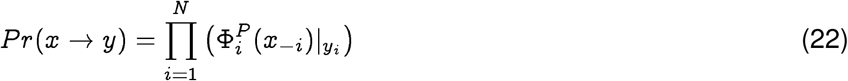

Writing the factors on the right hand side as:

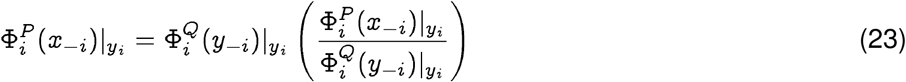

we note that 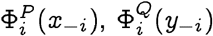, are distributions on the same index *i*, and hence:

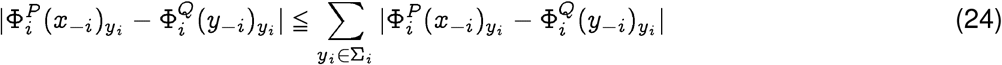

Using a standard refinement of Pinsker’s inequality ^9^, and the relationship of Jensen-Shannon divergence with total variation, we get:

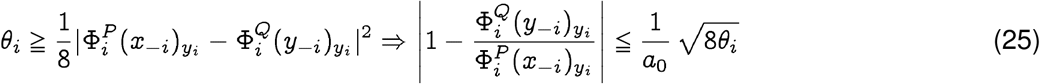

where *a*_0_ is the smallest non-zero probability value of generating the entry at any index. We will see that this parameter is related to statistical significance of our bounds. First, we can formulate a lower bound as follows:

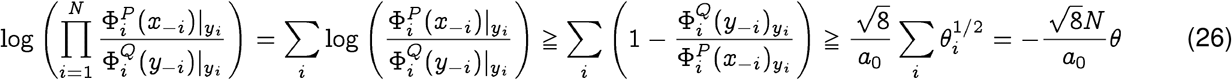

Similarly, the upper bound may be derived as:

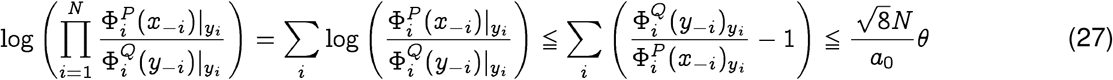

Combining Eqs. 26 and 27, we conclude:

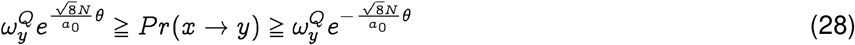

Now, interpreting *a*_0_ as the probability of generating an unlikely event below our desired threshold(*i.e*. a “failure”), we note that the probability of generating at least one such event is given by 1 − (1 − *a*_0_)^*N*^. Hence if *α* is the pre-specified significance level, we have for N >> 1:

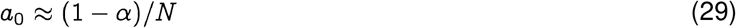

Hence, we conclude, that at significance level ≧*α*, we have the bounds:

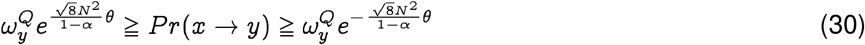

□

##### Remark 1.

*This bound can be rewritten in terms of the log-likelihood of the spontaneous jump and constants independent of the initial sequence x as:*

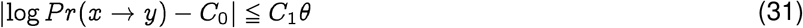

*where the constants are given by:*

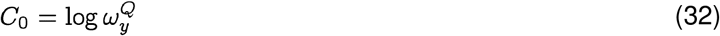

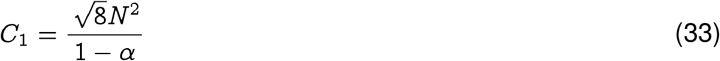

#### Multivariate Regression to Identify Factors in Strain Prediction

We investigate the key factors that contribute to our successful prediction of the dominant strain in the next season. We carry out a multivariate regression with data diversity, the complexity of inferred Qnet and the edit distance of the WHO recommendation from the dominant strain as independent variables. Here we define data diversity as the number of clusters we have in the input set of sequences, such that any two sequences five or less mutations apart are in the same cluster. Qnet complexity is measured by the number of decision nodes in the component decision trees of the recursive forest.

We select several plausible structures of the regression equation, and in each case conclude that data diversity has the most important and statistically significant contribution (See Tab. IV).

---

**Fig 1.**
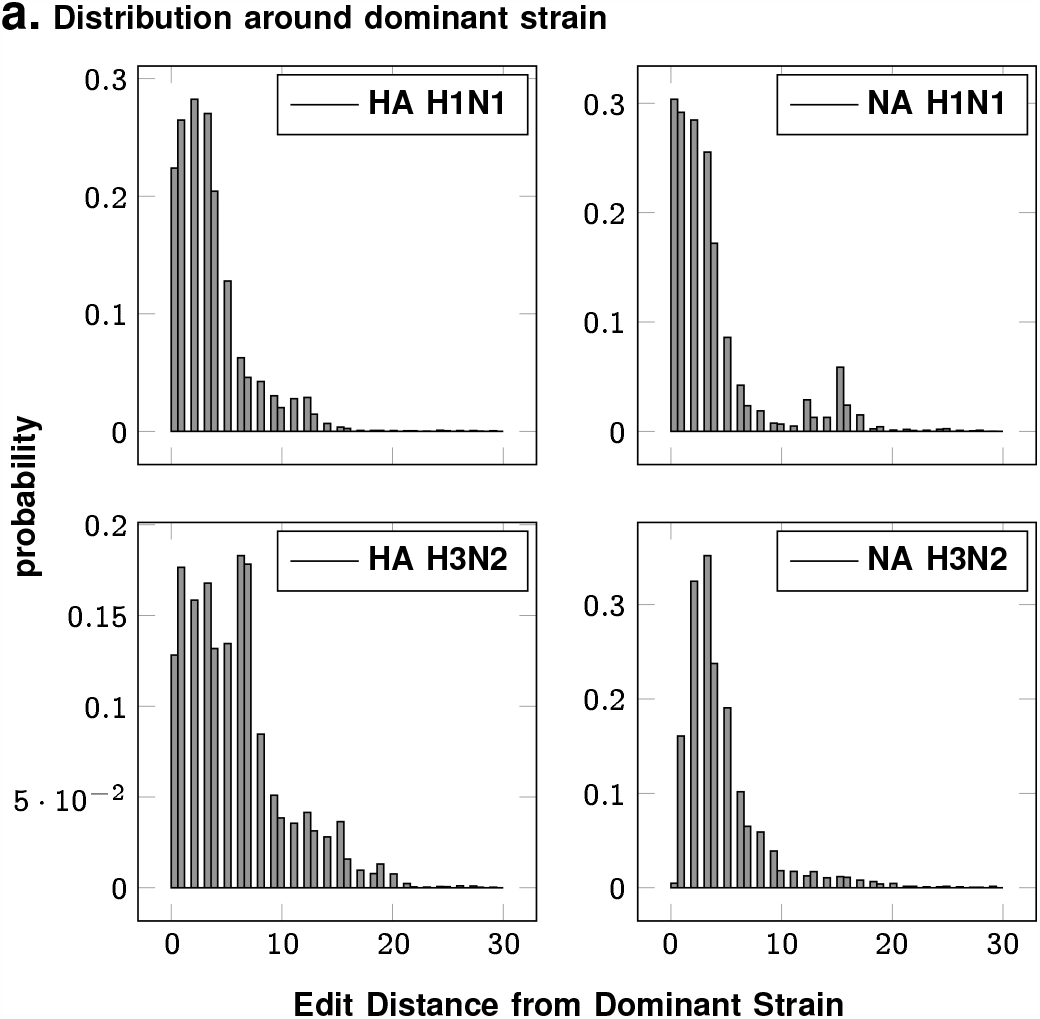
No. of mutations from the seasonal dominant strain over the years. The quasispecies that circulates each season for each subtype is tightly distributed around the dominant strain on average.

**TABLE I.** Eaxmples: Qnet induced distance varying for fixed sequence pair when background population changes (rows 1-5), sequences with small edit distance and large q-distance, and the converse (rows 6-9)

|  | edit dist. | sequence A | sequence B | q-distance | year A* | year B* |
| --- | --- | --- | --- | --- | --- | --- |
| 1 | 18 | A/Singapore/23J/2007 | A/Tennessee/UR06-0294/2007 | 0.0111 | 2007 | 2007 |
| 2 | 18 | A/Singapore/23J/2007 | A/Tennessee/UR06-0294/2007 | 0.0094 | 2008 | 2008 |
| 3 | 18 | A/Singapore/23J/2007 | A/Tennessee/UR06-0294/2007 | 0.0027 | 2009 | 2009 |
| 4 | 18 | A/Singapore/23J/2007 | A/Tennessee/UR06-0294/2007 | 0.0025 | 2010 | 2010 |
| 5 | 18 | A/Singapore/23J/2007 | A/Tennessee/UR06-0294/2007 | 0.6163 | 2007 | 2010 |
| 6 | 11 | A/Naypyitaw/M783/2008 | A/Singapore/201/2008 | 0.8852 | 2008 | 2008 |
| 7 | 15 | A/Cambodia/W0908339/2012 | A/Singapore/DMS1233/2012 | 0.2737 | 2012 | 2012 |
| 8 | 126 | A/South Dakota/03/2008 | A/Singapore/10/2008 | 0.3034 | 2008 | 2008 |
| 9 | 141 | A/Jodhpur/3248/2012 | A/Cambodia/W0908339/2012 | 0.2405 | 2012 | 2012 |

\*year A and year B correspond to the assumed collection years for sequences A and B respectively for the purpose of this example. Sequence A in row 1 is collected in 2007, but is assumed to be from different years in rows 2-4 to demonstrate the change in q-distance from sequence B, arising only from a change in the background population.

**TABLE II.** Correlation between q-distance and edit distance between sequence pairs

| phenotypes | correlation |
| --- | --- |
| Influenza H1N1 HA | 0.76 |
| Influenza H1N1 NA | 0.74 |
| Influenza H3N2 HA | 0.85 |
| Influenza H3N2 NA | 0.79 |
| SARS-CoV-2 | 0.52 |

**TABLE III.** Number of sequences collected from public databases

| Database | Strain | No. of Sequences |
| --- | --- | --- |
| NCBI | Influenza H1N1 HA | 7,761 |
| NCBI | Influenza H1N1 NA | 5,640 |
| NCBI | Influenza H3N2 HA | 6,568 |
| GISAID | Influenza H3N2 HA | 2,000 |
| NCBI | Influenza H3N2 NA | 4,919 |
| GISAID | Influenza H3N2 NA | 2,000 |
| NCBI | SARS-CoV-2 | 24 |
| GISAID | SARS-CoV-2 | 371 |
| NCBI | betacoronavirus (non-SARS-CoV-2) | 921 |
| Total |  | 30,204 |

**Fig 2.**
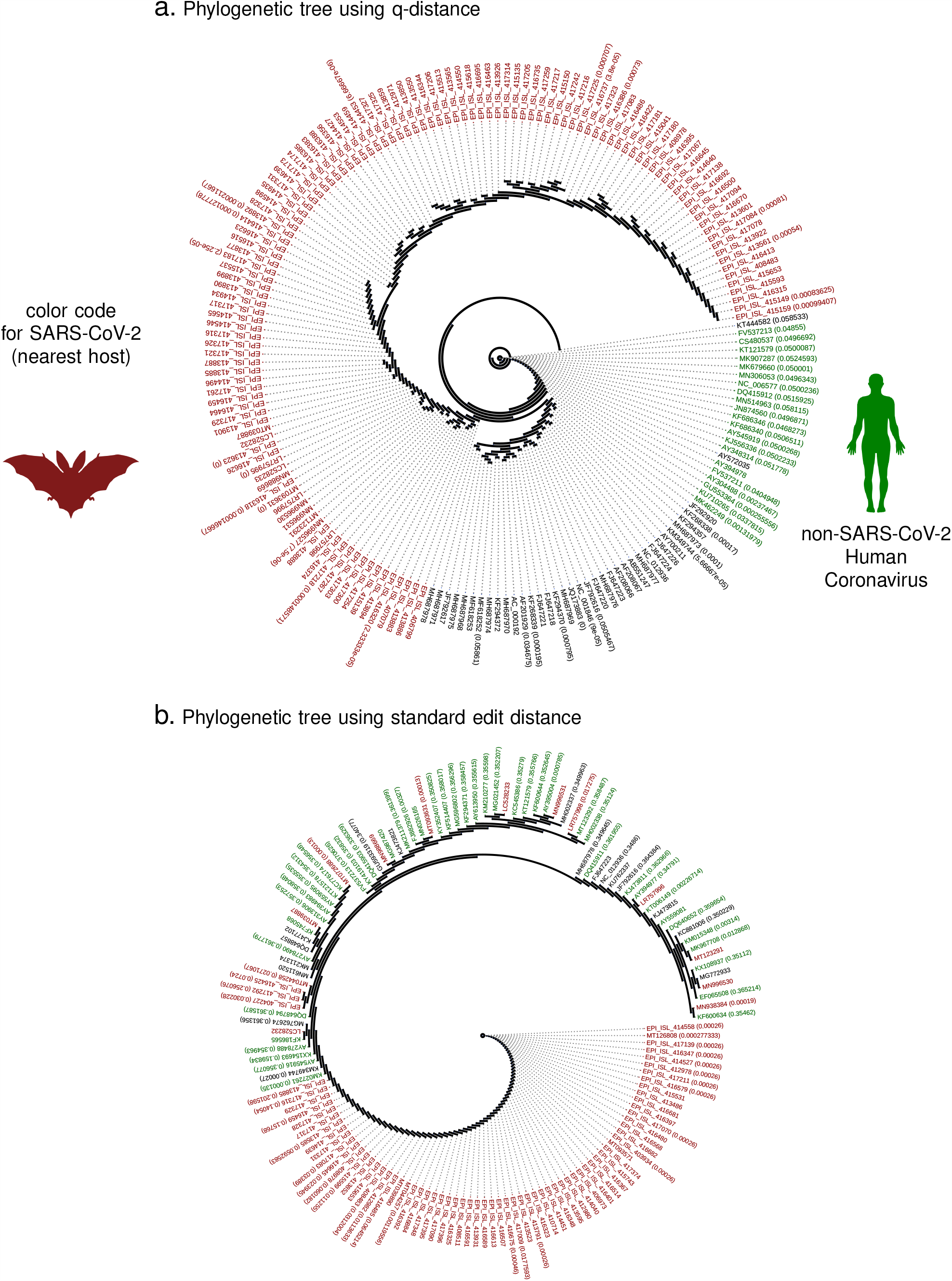
Phylogeny comparison between q-distance (panel a) and classical edit distance (panel b). The numbers within brackets is the distance within which the specific branch is collapsed for visualization. The classical distance produces a phylogeny which clearly violates chronological ordering, arising the novel coronavirus appears before strains that have been collected years before, including the SARs-1 strains. The new distance using Qnet is shown to automatically respect this known ordering.

**Fig 3.**
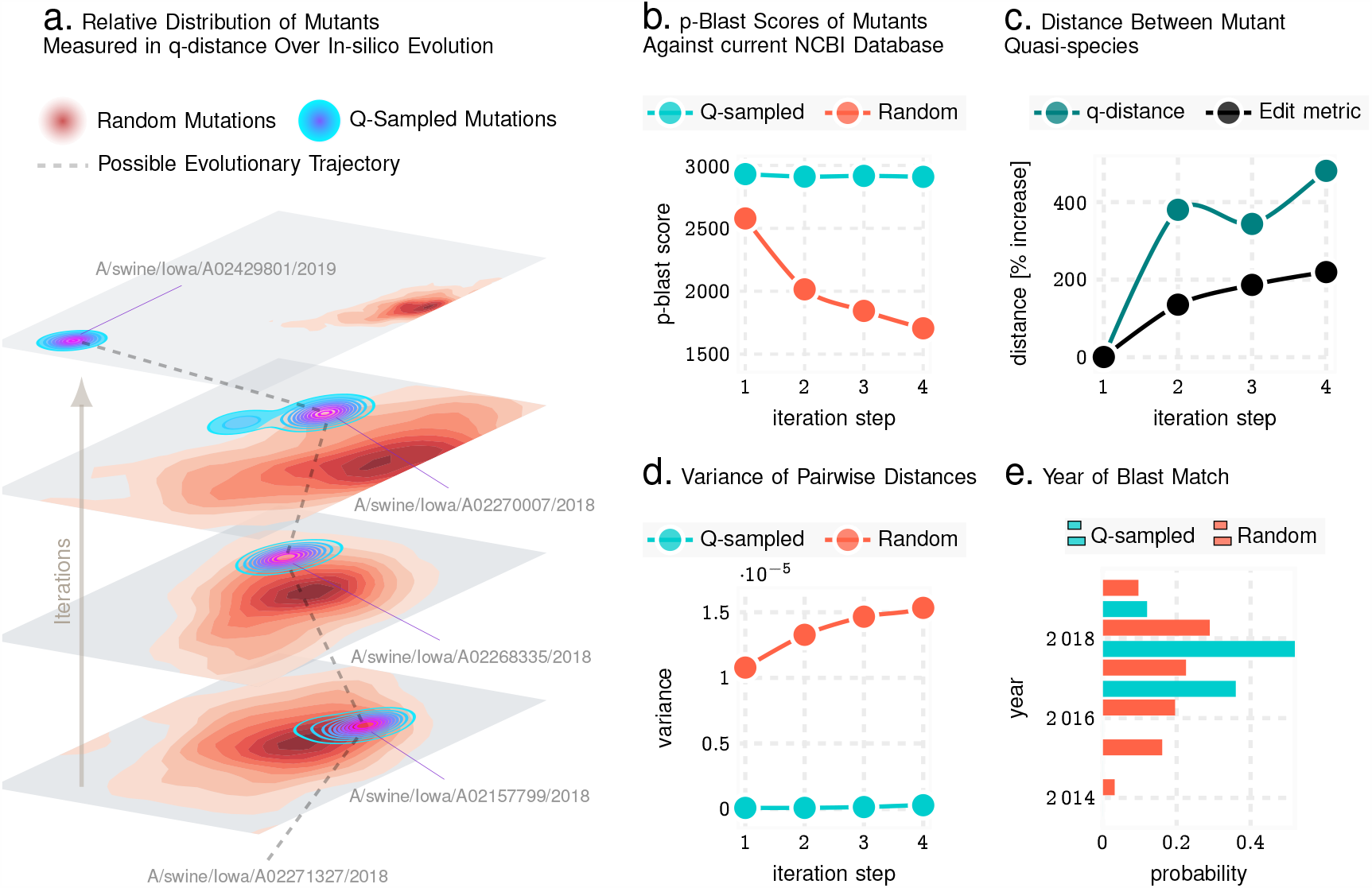
Q-distance validation in silico using Influenza A sequences from NCBI database. Panel a illustrates that the Qnet induced modeling of evolutionary trajectories initiated from known haemagglutinnin (HA) sequences are distinct from random paths in the strain space. In particular, random trajectories have more variance, and more importantly, diverge to different regions of the landscape compared to Qnet predictions. Panel b-e show that unconstrained Q-sampling produces sequences maintain a higher degree of similarity to known sequences, as verified by blasting against known HA sequences, have a smaller rate of growth of variance, and produce matches in closer time frames to the initial sequence. Panel c shows that this is not due to simply restricting the mutational variations, which increases rapidly in both the Qnet and the classical metric.

**Fig 4.**
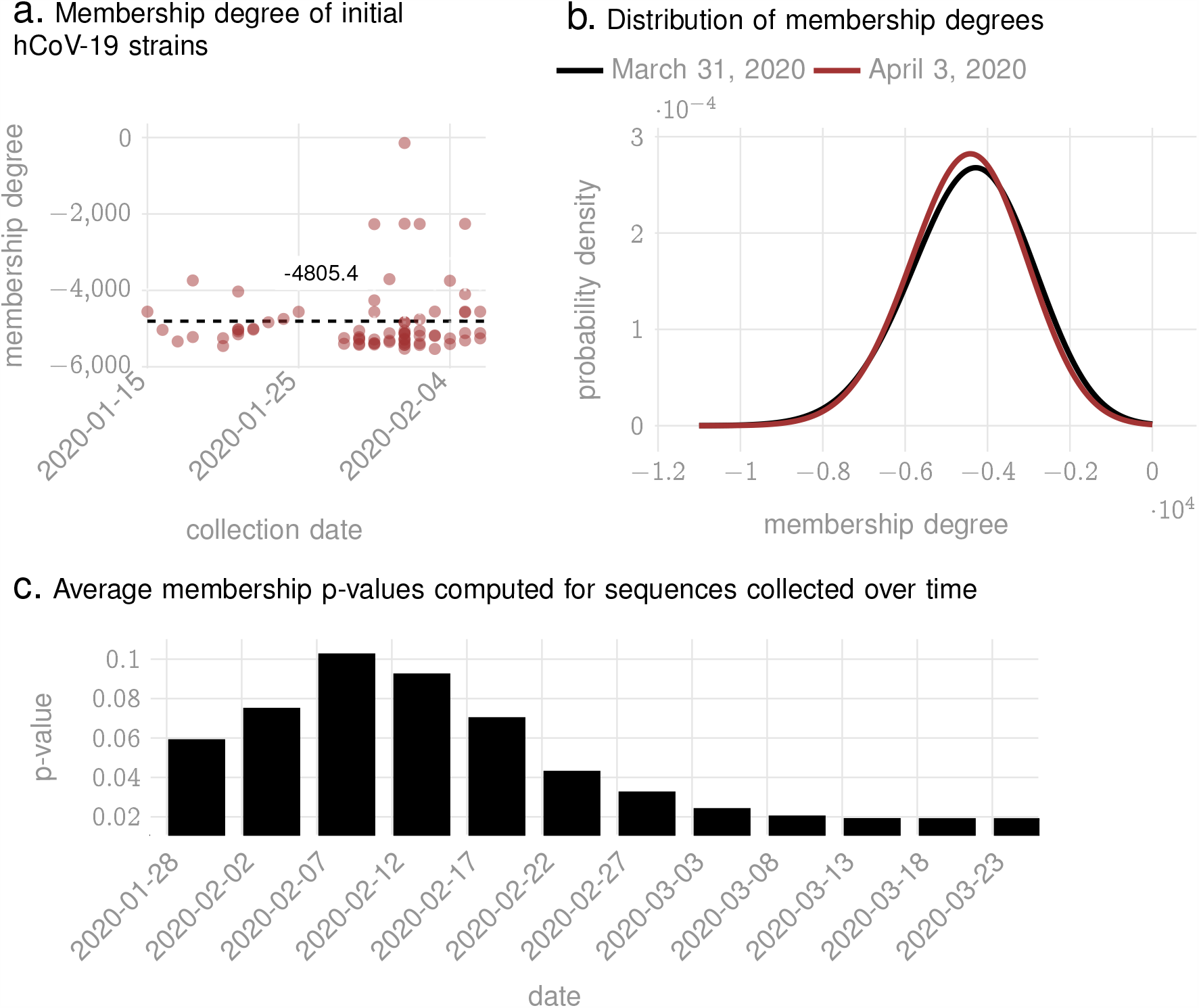
Membership degrees for SARS-CoV-2 sequences collected in the early days of the pandemic. The membership degree quantifies the likelihood that a test sequence actually is generated by the inferred model, i.e., the Qnet (See Methods for definition of membership degree).

**TABLE IV.** General linear model for evaluating effect of data diversity on Qnet performance

| variable name | description |
| --- | --- |
| qnet_complexity | Cumulative number of nodes in all predictors in the corresponding Qnet |
| data_diversity | Number of clusters in set of input sequence where each sequence in a specific cluster is separated by at least 5 mutations from sequences not in the cluster |
| ldistance_WHO | Deviation of WHO predicted strain from the dominant strain |

|  | coef | std err | z | P>\|z\| | [0.025 | 0.975] |
| --- | --- | --- | --- | --- | --- | --- |
| Intercept | -0.1116 | 1.090 | -0.102 | 0.918 | -2.248 | 2.025 |
| qnet_complexity | 0.0005 | 0.000 | 1.075 | 0.282 | -0.000 | 0.001 |
| data_diversity | 0.3197 | 0.126 | 2.531 | 0.011 | 0.072 | 0.567 |
| qnet_complexity:data_diversity | -6.932e-05 | 5.01e-05 | -1.383 | 0.167 | -0.000 | 2.89e-05 |
| ldistance_WHO | -0.0348 | 0.035 | -1.007 | 0.314 | -0.102 | 0.033 |

|  | coef | std err | z | P>\|z\| | [0.025 | 0.975] |
| --- | --- | --- | --- | --- | --- | --- |
| Intercept | 1.0841 | 0.665 | 1.630 | 0.103 | -0.219 | 2.387 |
| qnet_complexity | -4.12e-05 | 0.000 | -0.156 | 0.876 | -0.001 | 0.000 |
| data_diversity | 0.1788 | 0.075 | 2.392 | 0.017 | 0.032 | 0.325 |
| ldistance_WHO | -0.0695 | 0.024 | -2.930 | 0.003 | -0.116 | -0.023 |

**TABLE V.** H1N1 HA Northern Hemisphere

| year | WHO recommendation | dominant strain | Qnet recommendation | WHO error | Qnet error |
| --- | --- | --- | --- | --- | --- |
| 2001-2002 | A/New Caledonia/20/99 | A/Canterbury/41/2001 | A/Dunedin/2/2000 | 4 | 6 |
| 2002-2003 | A/New Caledonia/20/99 | A/Taiwan/567/2002 | A/Canterbury/41/2001 | 3 | 1 |
| 2003-2004 | A/New Caledonia/20/99 | A/Memphis/5/2003 | A/New York/291/2002 | 5 | 2 |
| 2004-2005 | A/New Caledonia/20/99 | A/Thailand/Siriraj-Rama-TT/2004 | A/Memphis/5/2003 | 7 | 4 |
| 2005-2006 | A/New Caledonia/20/99 | A/Niedersachsen/217/2005 | A/Canterbury/106/2004 | 8 | 10 |
| 2006-2007 | A/New Caledonia/20/99 | A/India/34980/2006 | A/Auckland/619/2005 | 6 | 1 |
| 2007-2008 | A/Solomon Islands/3/2006 | A/Norway/1701/2007 | A/Auckland/619/2005 | 8 | 11 |
| 2008-2009 | A/Brisbane/59/2007 | A/Pennsylvania/02/2008 | A/Kentucky/UR06-0476/2007 | 2 | 2 |
| 2009-2010 | A/Brisbane/59/2007 | A/Singapore/ON1060/2009 | A/Belem/241/2008 | 119 | 119 |
| 2010-2011 | A/California/7/2009 | A/England/01220740/2010 | A/Singapore/ON1060/2009 | 5 | 1 |
| 2011-2012 | A/California/7/2009 | A/Punjab/041/2011 | A/England/01220740/2010 | 7 | 2 |
| 2012-2013 | A/California/7/2009 | A/British Columbia/001/2012 | A/Punjab/041/2011 | 11 | 4 |
| 2013-2014 | A/California/7/2009 | A/Moscow/CRIE-32/2013 | A/Helsinki/1199/2012 | 10 | 2 |
| 2014-2015 | A/California/7/2009 | A/Thailand/CU-C5169/2014 | A/Thailand/CU-C5169/2014 | 12 | 0 |
| 2015-2016 | A/California/7/2009 | A/Georgia/15/2015 | A/Thailand/CU-C5169/2014 | 14 | 2 |
| 2016-2017 | A/California/7/2009 | A/Hawaii/21/2016 | A/Hawaii/21/2016 | 16 | 0 |
| 2017-2018 | A/Michigan/45/2015 | A/Michigan/291/2017 | A/Beijing-Huairou/SWL1335/2016 | 5 | 4 |
| 2018-2019 | A/Michigan/45/2015 | A/Washington/55/2018 | A/Michigan/291/2017 | 6 | 1 |
| 2019-2020 | A/Brisbane/02/2018 | A/Kentucky/06/2019 | A/Washington/55/2018 | 5 | 1 |
| 2020-2021 | A/Hawaii/70/2019 | -1 | A/Italy/8451/2019 | -1 | -1 |

\* Dominant strain is calculated as the one closest to the centroid in the strain space that year in the edit distance metric

**TABLE VI.**
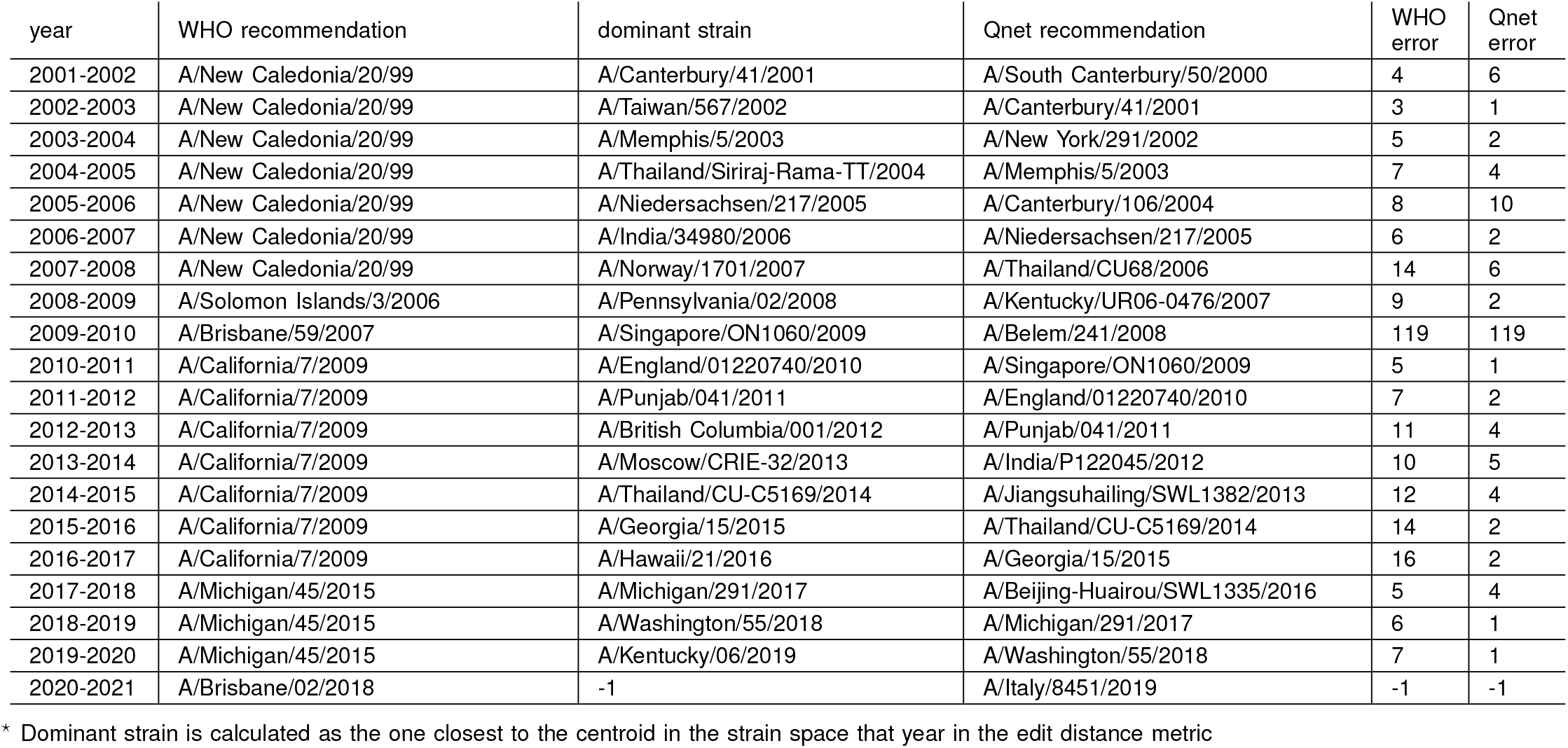
H1N1 HA Southern Hemisphere

**TABLE VII.**
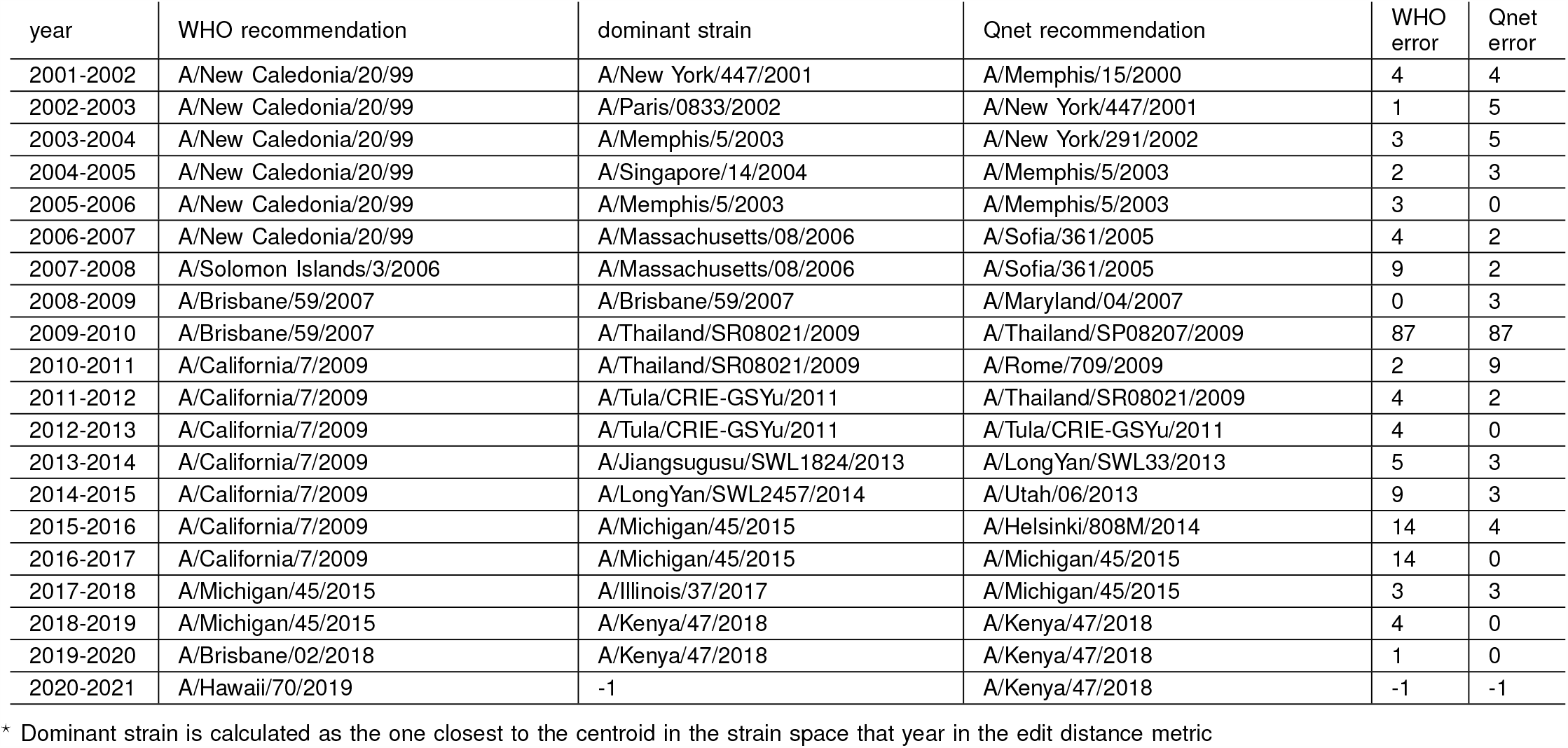
H1N1 NA Northern Hemisphere

**TABLE VIII.**
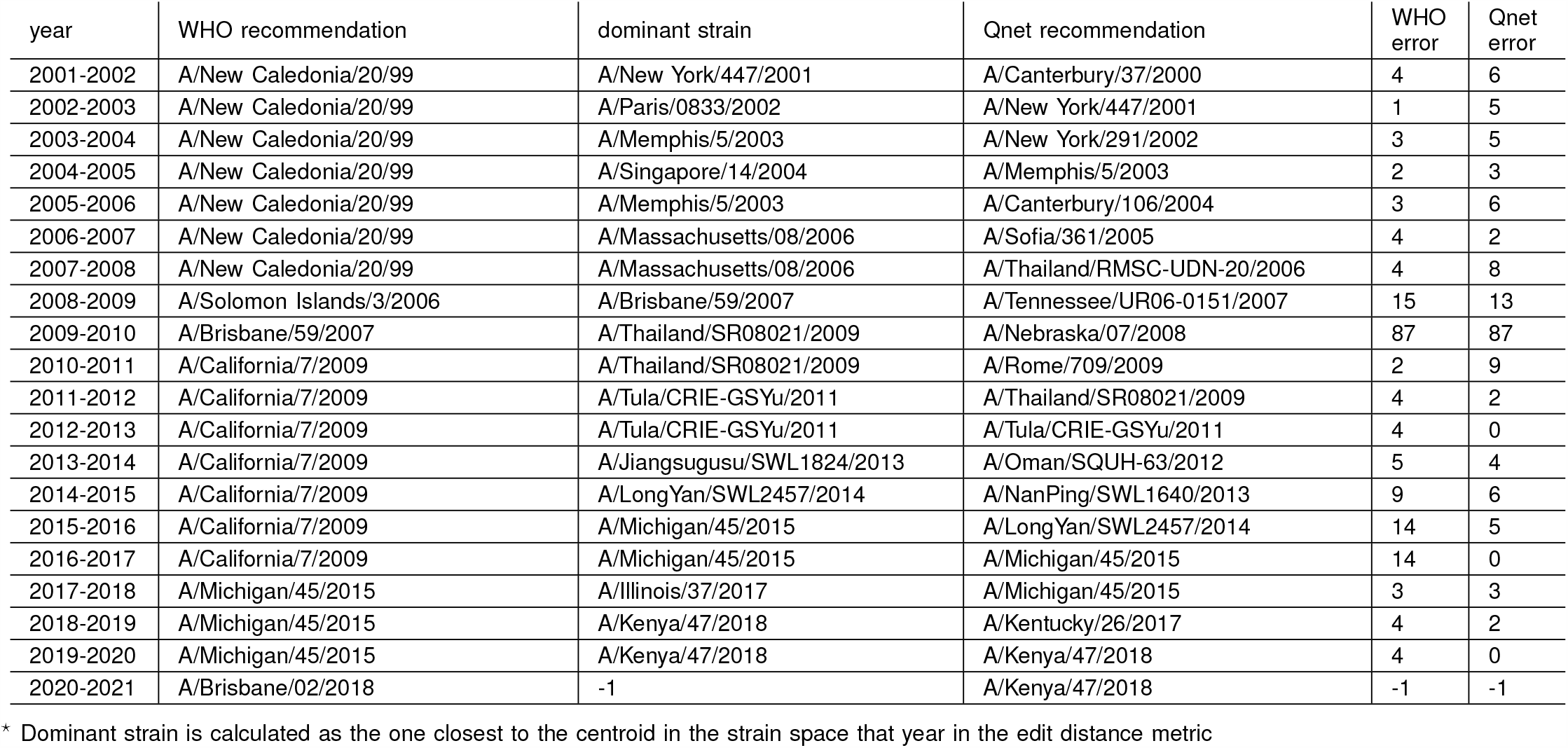
H1N1 NA Southern Hemisphere

| year | WHO recommendation | dominant strain | Qnet recommendation | WHO error | Qnet error |
| --- | --- | --- | --- | --- | --- |
| 2001-2002 | A/New Caledonia/20/99 | A/New York/447/2001 | A/Canterbury/37/2000 | 4 | 6 |
| 2002-2003 | A/New Caledonia/20/99 | A/Paris/0833/2002 | A/New York/447/2001 | 1 | 5 |
| 2003-2004 | A/New Caledonia/20/99 | A/Memphis/5/2003 | A/New York/291/2002 | 3 | 5 |
| 2004-2005 | A/New Caledonia/20/99 | A/Singapore/14/2004 | A/Memphis/5/2003 | 2 | 3 |
| 2005-2006 | A/New Caledonia/20/99 | A/Memphis/5/2003 | A/Canterbury/106/2004 | 3 | 6 |
| 2006-2007 | A/New Caledonia/20/99 | A/Massachusetts/08/2006 | A/Sofia/361/2005 | 4 | 2 |
| 2007-2008 | A/New Caledonia/20/99 | A/Massachusetts/08/2006 | A/Thailand/RMSC-UDN-20/2006 | 4 | 8 |
| 2008-2009 | A/Solomon Islands/3/2006 | A/Brisbane/59/2007 | A/Tennessee/UR06-0151/2007 | 15 | 13 |
| 2009-2010 | A/Brisbane/59/2007 | A/Thailand/SR08021/2009 | A/Nebraska/07/2008 | 87 | 87 |
| 2010-2011 | A/California/7/2009 | A/Thailand/SR08021/2009 | A/Rome/709/2009 | 2 | 9 |
| 2011-2012 | A/California/7/2009 | A/Tula/CRIE-GSYu/2011 | A/Thailand/SR08021/2009 | 4 | 2 |
| 2012-2013 | A/California/7/2009 | A/Tula/CRIE-GSYu/2011 | A/Tula/CRIE-GSYu/2011 | 4 | 0 |
| 2013-2014 | A/California/7/2009 | A/Jiangsugusu/SWL1824/2013 | A/Oman/SQUH-63/2012 | 5 | 4 |
| 2014-2015 | A/California/7/2009 | A/LongYan/SWL2457/2014 | A/NanPing/SWL1640/2013 | 9 | 6 |
| 2015-2016 | A/California/7/2009 | A/Michigan/45/2015 | A/LongYan/SWL2457/2014 | 14 | 5 |
| 2016-2017 | A/California/7/2009 | A/Michigan/45/2015 | A/Michigan/45/2015 | 14 | 0 |
| 2017-2018 | A/Michigan/45/2015 | A/Illinois/37/2017 | A/Michigan/45/2015 | 3 | 3 |
| 2018-2019 | A/Michigan/45/2015 | A/Kenya/47/2018 | A/Kentucky/26/2017 | 4 | 2 |
| 2019-2020 | A/Michigan/45/2015 | A/Kenya/47/2018 | A/Kenya/47/2018 | 4 | 0 |
| 2020-2021 | A/Brisbane/02/2018 | -1 | A/Kenya/47/2018 | -1 | -1 |

\* Dominant strain is calculated as the one closest to the centroid in the strain space that year in the edit distance metric

**TABLE IX.** H3N2 HA Northern Hemisphere

| year | WHO recommendation | dominant strain | Qnet recommendation | WHO error | Qnet error |
| --- | --- | --- | --- | --- | --- |
| 2005-2006 | A/California/7/2004 | A/Denmark/195/2005 | A/Tairawhiti/369/2004 | 10 | 2 |
| 2006-2007 | A/Wisconsin/67/2005 | A/New York/5/2006 | A/South Australia/22/2005 | 5 | 4 |
| 2007-2008 | A/Wisconsin/67/2005 | A/Tennessee/11/2007 | A/Colorado/05/2006 | 8 | 5 |
| 2008-2009 | A/Brisbane/10/2007 | A/Massachusetts/13/2008 | A/Tennessee/11/2007 | 3 | 2 |
| 2009-2010 | A/Brisbane/10/2007 | A/Hawaii/14/2009 | A/Manhean/03/2008 | 7 | 6 |
| 2010-2011 | A/Perth/16/2009 | A/Utah/12/2010 | A/Hawaii/14/2009 | 8 | 7 |
| 2011-2012 | A/Perth/16/2009 | A/Piaui/14202/2011 | A/Utah/12/2010 | 4 | 4 |
| 2012-2013 | A/Victoria/361/2011 | A/Alborz/927/2012 | A/Tehran/895/2012 | 4 | 3 |
| 2013-2014 | A/Victoria/361/2011 | A/Delaware/01/2013 | A/Singapore/H2012.934/2012 | 4 | 1 |
| 2014-2015 | A/Texas/50/2012 | A/Hong Kong/4801/2014 | A/Nebraska/03/2013 | 10 | 9 |
| 2015-2016 | A/Switzerland/9715293/2013 | A/Hong Kong/4801/2014 | A/Hong Kong/4801/2014 | 10 | 0 |
| 2016-2017 | A/Hong Kong/4801/2014 | A/Hong Kong/4801/2014 | A/Hong Kong/4801/2014 | 0 | 0 |
| 2017-2018 | A/Hong Kong/4801/2014 | A/Maryland/25/2017 | A/New York/03/2016 | 3 | 1 |
| 2018-2019 | A/Singapore/INFIMH-16-0019/2016 | A/Vermont/04/2018 | A/Ontario/038/2017 | 8 | 5 |
| 2019-2020 | A/Kansas/14/2017 | A/Kentucky/27/2019 | A/California/7330/2018 | 16 | 12 |
| 2020-2021 | A/Hong Kong/2671/2019 | -1 | A/Kentucky/27/2019 | -1 | -1 |

\* Dominant strain is calculated as the one closest to the centroid in the strain space that year in the edit distance metric

**TABLE X.** H3N2 HA Southern Hemisphere

| year | WHO recommendation | dominant strain | Qnet recommendation | WHO error | Qnet error |
| --- | --- | --- | --- | --- | --- |
| 2005-2006 | A/Wellington/1/2004 | A/Denmark/195/2005 | A/Waikato/21/2004 | 3 | 3 |
| 2006-2007 | A/California/7/2004 | A/New York/5/2006 | A/South Australia/22/2005 | 12 | 4 |
| 2007-2008 | A/Wisconsin/67/2005 | A/Tennessee/11/2007 | A/New York/923/2006 | 8 | 5 |
| 2008-2009 | A/Brisbane/10/2007 | A/Massachusetts/13/2008 | A/Tennessee/11/2007 | 3 | 2 |
| 2009-2010 | A/Brisbane/10/2007 | A/Hawaii/14/2009 | A/Manhean/03/2008 | 7 | 6 |
| 2010-2011 | A/Perth/16/2009 | A/Utah/12/2010 | A/Hawaii/14/2009 | 8 | 7 |
| 2011-2012 | A/Perth/16/2009 | A/Piaui/14202/2011 | A/Utah/12/2010 | 4 | 4 |
| 2012-2013 | A/Perth/16/2009 | A/Alborz/927/2012 | A/Piaui/14202/2011 | 8 | 4 |
| 2013-2014 | A/Victoria/361/2011 | A/Delaware/01/2013 | A/Callao/IPE00830/2012 | 4 | 7 |
| 2014-2015 | A/Texas/50/2012 | A/Hong Kong/4801/2014 | A/Delaware/01/2013 | 10 | 7 |
| 2015-2016 | A/Switzerland/9715293/2013 | A/Hong Kong/4801/2014 | A/Hong Kong/4801/2014 | 10 | 0 |
| 2016-2017 | A/Hong Kong/4801/2014 | A/Hong Kong/4801/2014 | A/Hong Kong/4801/2014 | 0 | 0 |
| 2017-2018 | A/Hong Kong/4801/2014 | A/Maryland/25/2017 | A/Ontario/196/2016 | 3 | 4 |
| 2018-2019 | A/Singapore/INFIMH-16-0019/2016 | A/Vermont/04/2018 | A/Ontario/038/2017 | 8 | 5 |
| 2019-2020 | A/Switzerland/8060/2017 | A/Kentucky/27/2019 | A/California/7330/2018 | 13 | 12 |
| 2020-2021 | A/South Australia/34/2019 | -1 | A/Kentucky/27/2019 | -1 | -1 |

\* Dominant strain is calculated as the one closest to the centroid in the strain space that year in the edit distance metric

**TABLE XI.**
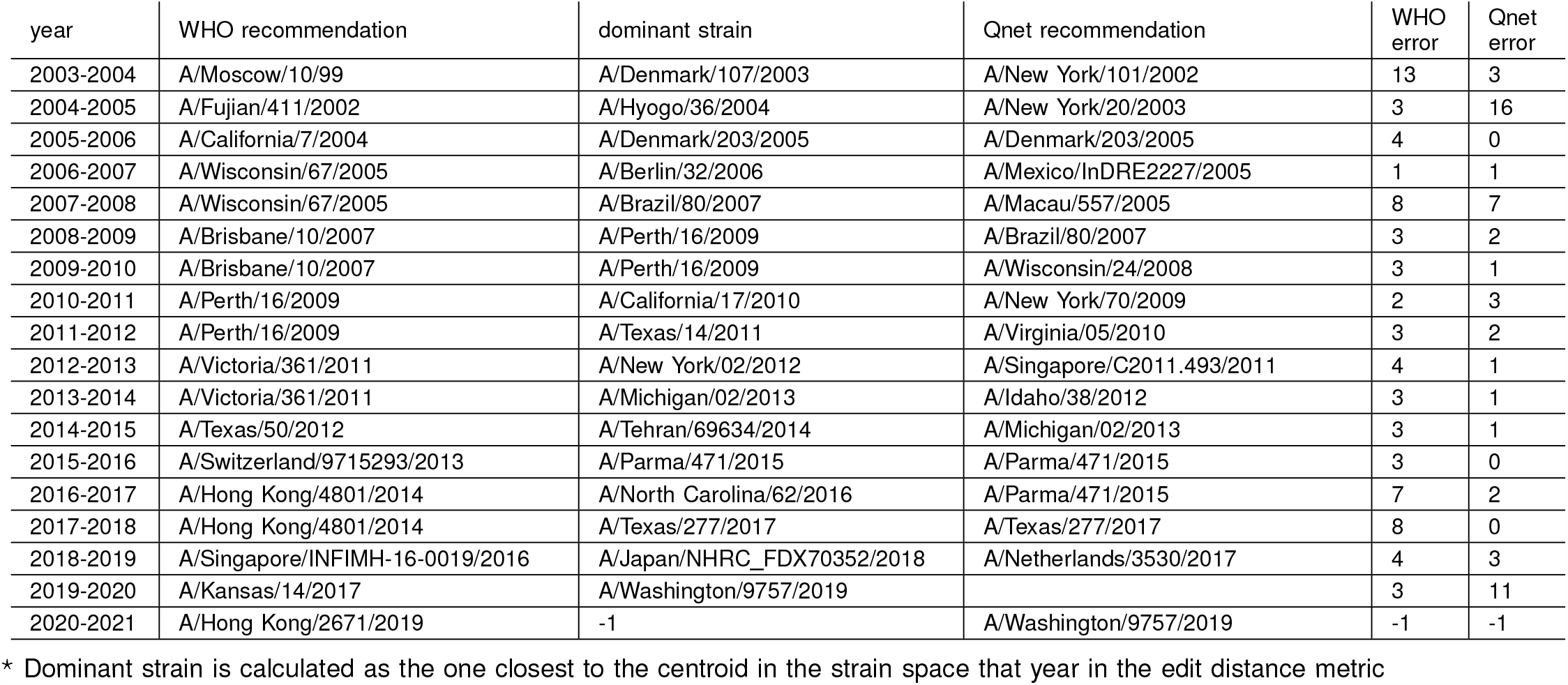
H3N2 NA Northern Hemisphere

| year | WHO recommendation | dominant strain | Qnet recommendation | WHO error | Qnet error |
| --- | --- | --- | --- | --- | --- |
| 2003-2004 | A/Moscow/10/99 | A/Denmark/107/2003 | A/New York/101/2002 | 13 | 3 |
| 2004-2005 | A/Fujian/411/2002 | A/Hyogo/36/2004 | A/New York/20/2003 | 3 | 16 |
| 2005-2006 | A/California/7/2004 | A/Denmark/203/2005 | A/Denmark/203/2005 | 4 | 0 |
| 2006-2007 | A/Wisconsin/67/2005 | A/Berlin/32/2006 | A/Mexico/InDRE2227/2005 | 1 | 1 |
| 2007-2008 | A/Wisconsin/67/2005 | A/Brazil/80/2007 | A/Macau/557/2005 | 8 | 7 |
| 2008-2009 | A/Brisbane/10/2007 | A/Perth/16/2009 | A/Brazil/80/2007 | 3 | 2 |
| 2009-2010 | A/Brisbane/10/2007 | A/Perth/16/2009 | A/Wisconsin/24/2008 | 3 | 1 |
| 2010-2011 | A/Perth/16/2009 | A/California/17/2010 | A/New York/70/2009 | 2 | 3 |
| 2011-2012 | A/Perth/16/2009 | A/Texas/14/2011 | A/Virginia/05/2010 | 3 | 2 |
| 2012-2013 | A/Victoria/361/2011 | A/New York/02/2012 | A/Singapore/C2011.493/2011 | 4 | 1 |
| 2013-2014 | A/Victoria/361/2011 | A/Michigan/02/2013 | A/Idaho/38/2012 | 3 | 1 |
| 2014-2015 | A/Texas/50/2012 | A/Tehran/69634/2014 | A/Michigan/02/2013 | 3 | 1 |
| 2015-2016 | A/Switzerland/9715293/2013 | A/Parma/471/2015 | A/Parma/471/2015 | 3 | 0 |
| 2016-2017 | A/Hong Kong/4801/2014 | A/North Carolina/62/2016 | A/Parma/471/2015 | 7 | 2 |
| 2017-2018 | A/Hong Kong/4801/2014 | A/Texas/277/2017 | A/Texas/277/2017 | 8 | 0 |
| 2018-2019 | A/Singapore/INFIMH-16-0019/2016 | A/Japan/NHRC_FDX70352/2018 | A/Netherlands/3530/2017 | 4 | 3 |
| 2019-2020 | A/Kansas/14/2017 | A/Washington/9757/2019 |  | 3 | 11 |
| 2020-2021 | A/Hong Kong/2671/2019 | -1 | A/Washington/9757/2019 | -1 | -1 |

\* Dominant strain is calculated as the one closest to the centroid in the strain space that year in the edit distance metric

**TABLE XII.** H3N2 NA Southern Hemisphere

| year | WHO recommendation | dominant strain | Qnet recommendation | WHO error | Qnet error |
| --- | --- | --- | --- | --- | --- |
| 2003-2004 | A/Moscow/10/99 | A/Denmark/107/2003 | A/New York/101/2002 | 13 | 3 |
| 2004-2005 | A/Fujian/411/2002 | A/Hyogo/36/2004 | A/New York/20/2003 | 3 | 16 |
| 2005-2006 | A/Wellington/1/2004 | A/Denmark/203/2005 | A/Wellington/1/2004 | 2 | 2 |
| 2006-2007 | A/California/7/2004 | A/Berlin/32/2006 | A/Mexico/InDRE2227/2005 | 3 | 1 |
| 2007-2008 | A/Wisconsin/67/2005 | A/Brazil/80/2007 | A/Ohio/06/2006 | 8 | 10 |
| 2008-2009 | A/Brisbane/10/2007 | A/Perth/16/2009 | A/Brazil/80/2007 | 3 | 2 |
| 2009-2010 | A/Brisbane/10/2007 | A/Perth/16/2009 | A/Wisconsin/24/2008 | 3 | 1 |
| 2010-2011 | A/Perth/16/2009 | A/California/17/2010 | A/New York/70/2009 | 2 | 3 |
| 2011-2012 | A/Perth/16/2009 | A/Texas/14/2011 | A/Virginia/05/2010 | 3 | 2 |
| 2012-2013 | A/Perth/16/2009 | A/New York/02/2012 | A/Texas/14/2011 | 4 | 1 |
| 2013-2014 | A/Victoria/361/2011 | A/Michigan/02/2013 | A/New York/02/2012 | 3 | 3 |
| 2014-2015 | A/Texas/50/2012 | A/Tehran/69634/2014 | A/Michigan/02/2013 | 3 | 1 |
| 2015-2016 | A/Switzerland/9715293/2013 | A/Parma/471/2015 | A/Tehran/69634/2014 | 3 | 2 |
| 2016-2017 | A/Hong Kong/4801/2014 | A/North Carolina/62/2016 | A/Parma/471/2015 | 7 | 2 |
| 2017-2018 | A/Hong Kong/4801/2014 | A/Texas/277/2017 | A/Texas/277/2017 | 8 | 0 |
| 2018-2019 | A/Singapore/INFIMH-16-0019/2016 | A/Japan/NHRC_FDX70352/2018 | A/Texas/277/2017 | 4 | 3 |
| 2019-2020 | A/Switzerland/8060/2017 | A/Washington/9757/2019 | A/Pennsylvania/317/2018 | 10 | 10 |
| 2020-2021 | A/South Australia/34/2019 | -1 | A/Washington/9757/2019 | -1 | -1 |

\* Dominant strain is calculated as the one closest to the centroid in the strain space that year in the edit distance metric

**TABLE XIII.**
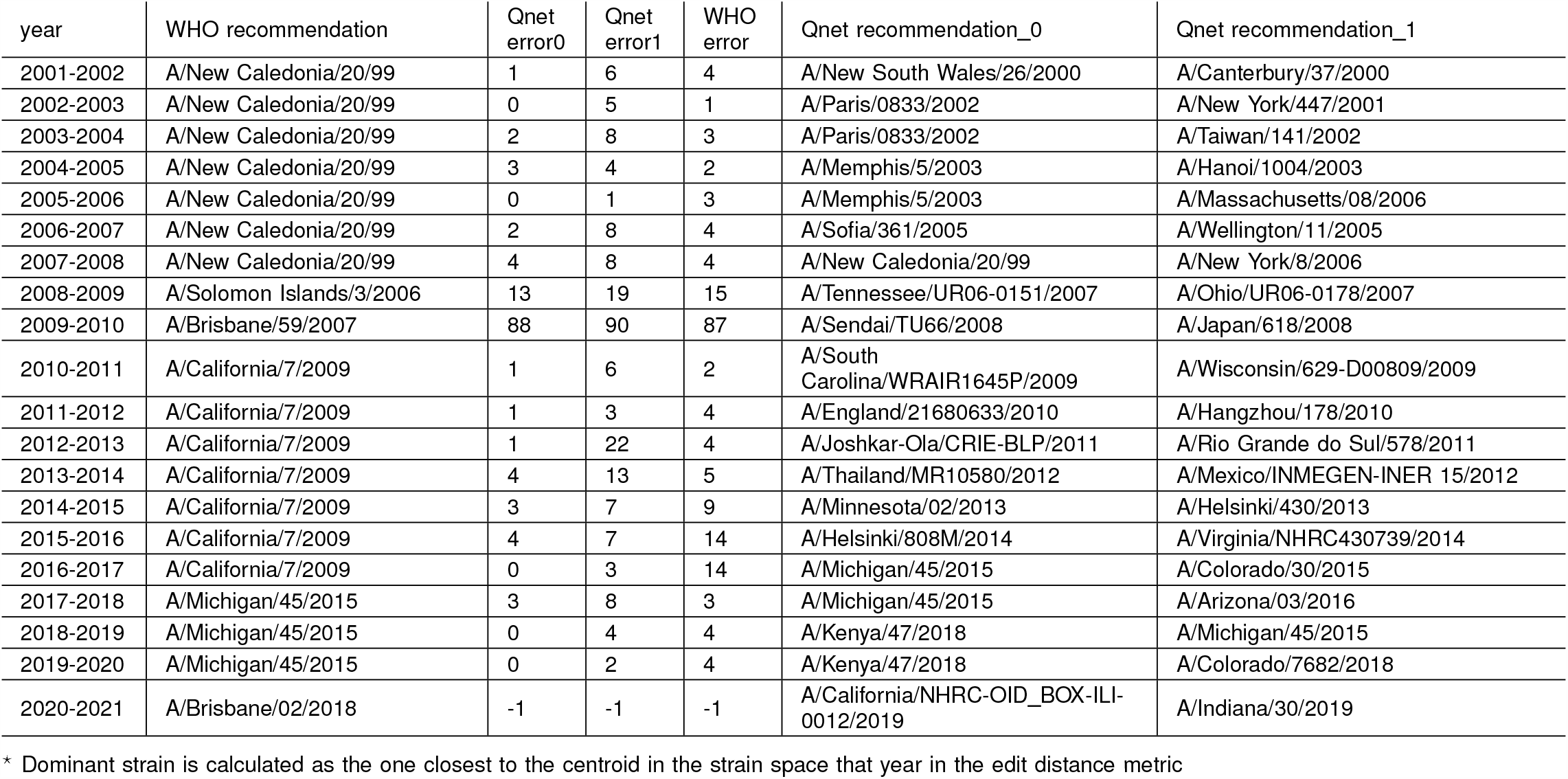
H1N1 NA Southern Hemisphere (Multi-cluster)

**TABLE XIV.** H3N2 NA Southern Hemisphere (Multi-cluster)

| year | WHO recommendation | Qnet<br>error0 | Qnet<br>error1 | WHO<br>error | Qnet recommendation_0 | Qnet recommendation_1 |
| --- | --- | --- | --- | --- | --- | --- |
| 2003-2004 | A/Moscow/10/99 | 4 | 5 | 13 | A/Auckland/612/2002 | A/New York/87/2002 |
| 2004-2005 | A/Fujian/411/2002 | 16 | 18 | 3 | A/New York/20/2003 | A/New York/12/2003 |
| 2005-2006 | A/Wellington/1/2004 | 1 | 7 | 2 | A/New York/358/2004 | A/Singapore/36/2004 |
| 2006-2007 | A/California/7/2004 | 3 | 8 | 3 | A/Macau/557/2005 | A/Hong Kong/HKU53/2005 |
| 2007-2008 | A/Wisconsin/67/2005 | 0 | 10 | 8 | A/Brazil/80/2007 | A/Wisconsin/44/2006 |
| 2008-2009 | A/Brisbane/10/2007 | 4 | 10 | 3 | A/Missouri/06/2007 | A/Japan/72/2007 |
| 2009-2010 | A/Brisbane/10/2007 | 1 | 7 | 3 | A/Wisconsin/24/2008 | A/Mississippi/UR07-0042/2008 |
| 2010-2011 | A/Perth/16/2009 | 3 | 8 | 2 | A/New York/70/2009 | A/Japan/883/2009 |
| 2011-2012 | A/Perth/16/2009 | 2 | 2 | 3 | A/California/19/2010 | A/Virginia/05/2010 |
| 2012-2013 | A/Perth/16/2009 | 1 | 12 | 4 | A/Texas/14/2011 | A/Singapore/GP1684/2011 |
| 2013-2014 | A/Victoria/361/2011 | 1 | 5 | 3 | A/Idaho/38/2012 | A/Pavia/135/2012 |
| 2014-2015 | A/Texas/50/2012 | 1 | 1 | 3 | A/Nevada/05/2013 | A/Michigan/02/2013 |
| 2015-2016 | A/Switzerland/9715293/2013 | 0 | 4 | 3 | A/Parma/471/2015 | A/Iran/91244/2014 |
| 2016-2017 | A/Hong Kong/4801/2014 | 1 | 25 | 7 | A/New Jersey/13/2015 | A/California/NHRC_BRD41056N/2015 |
| 2017-2018 | A/Hong Kong/4801/2014 | 1 | 4 | 9 | A/Texas/277/2017 | A/Victoria/668/2016 |
| 2018-2019 | A/Singapore/INFIMH-16-0019/2016 | 2 | 4 | 3 | A/Netherlands/3530/2017 | A/Washington/17/2017 |
| 2019-2020 | A/Switzerland/8060/2017 | 4 | 10 | 10 | A/England/538/2018 | A/California/BRD12490N/2018 |
| 2020-2021 | A/South Australia/34/2019 | -1 | -1 | -1 | A/South Australia/34/2019 | A/Washington/9757/2019 |

\* Dominant strain is calculated as the one closest to the centroid in the strain space that year in the edit distance metric

**TABLE XV.** Neighbors at the edge of emergence

| accession | country | date | qdistance* | host | log-likelihood bound† |
| --- | --- | --- | --- | --- | --- |
| MG197717 | China | 2015-07-06 | 0.5994 | human(Human coronavirus OC43) | -344680279.6919 |
| MG197719 | China | 2015-06-04 | 0.5995 | human(Human coronavirus OC43) | -344700037.5225 |
| MG197710 | China | 2015-05-06 | 0.6002 | human(Human coronavirus OC43) | -345145082.3718 |
| MH940245 | Thailand | 2017-06-04 | 0.6017 | human(Human coronavirus HKU1) | -346005398.0759 |
| MG197711 | China | 2015-06-09 | 0.6035 | human(Human coronavirus OC43) | -347017496.9155 |
| MG197716 | China | 2015-06-06 | 0.6053 | human(Human coronavirus OC43) | -348034981.4953 |
| MG197715 | China | 2015-05-21 | 0.6053 | human(Human coronavirus OC43) | -348055196.8970 |
| KF294457 | China | 2012-01-01 | 0.6058 | Rhinolophus monoceros | -348315726.8189 |
| KJ473822 | China | 2012-01-01 | 0.6059 | Tylonycteris pachypus | -348379385.2394 |
| MK211376 | China | 2016-09-01 | 0.6065 | Rhinolophus affinis | -348745110.7506 |
| MH002342 | China | 2013-06-03 | 0.6065 | Pipistrellus bat coronavirus HKU5 | -348745431.2254 |
| KJ473816 | China | 2013-01-01 | 0.6066 | Rhinolophus sinicus | -348779627.4654 |
| KJ473812 | China | 2013-01-01 | 0.6066 | Rhinolophus ferrumequinum | -348807413.3783 |
| MG772933 | China | 2017-02-01 | 0.6066 | Rhinolophus sinicus | -348814549.9518 |
| MK211379 | China | 2016-09-01 | 0.6067 | Rhinolophus affinis | -348846490.8570 |
| MK211375 | China | 2016-09-01 | 0.6067 | Rhinolophus affinis | -348867989.3104 |
| MK211374 | China | 2016-08-01 | 0.6068 | Rhinolophus sp. | -348893681.6418 |
| KJ473821 | China | 2014-05-06 | 0.6071 | Vespertilio superans | -349070089.5700 |
| KF569996 | China | 2011-01-01 | 0.6095 | Rhinolophus affinis | -350440764.5785 |
| MN611520 | China | 2018-03-01 | 0.6095 | Pipistrellus abramus | -350452309.6142 |
| KP886809 | China | 2013-05-23 | 0.6095 | Rhinolophus Ferrumequinum | -350486988.0164 |
| KP886808 | China | 2013-05-23 | 0.6095 | Rhinolophus Ferrumequinum | -350486988.0164 |
| MN611519 | China | 2018-03-01 | 0.6097 | Tylonycteris pachypus | -350572065.7797 |
| NC_025217 | China | 2013-04-29 | 0.6097 | Hipposideros pratti | -350580907.8832 |
| MK211377 | China | 2016-09-01 | 0.6106 | Rhinolophus affinis | -351127765.1568 |
| KJ473820 | China | 2013-01-01 | 0.6118 | Pipistrellus abramus | -351798100.8827 |
| MN996532‡ | China | 2013-07-24 | 0.6155 | Rhinolophus affinis | -353944009.5536 |
| MH002341 | China | 2014-06-28 | 0.6167 | Pipistrellus bat coronavirus HKU5 | -354632651.3696 |
| MH687968 | Viet Nam | 2014-11-14 | 0.6174 | Rattus argentiventer | -355004271.8441 |
| MH687978 | Viet Nam | 2015-02-04 | 0.6183 | Rattus argentiventer | -355553631.3715 |
| MH687969 | Viet Nam | 2014-11-12 | 0.6184 | Rattus argentiventer | -355566733.0149 |
| KF294372 | China | 2011-01-01 | 0.6185 | Niviventer confucianus | -355664120.7144 |
| MH687974 | Viet Nam | 2014-11-12 | 0.6187 | Rattus argentiventer | -355732457.2680 |
| MH687973 | Viet Nam | 2014-11-12 | 0.6189 | Rattus argentiventer | -355892765.0568 |
| MH687972 | Viet Nam | 2014-11-12 | 0.6190 | Rattus argentiventer | -355956649.5930 |
| KF294370 | China | 2013-01-01 | 0.6192 | Rattus tanezumi | -356024166.0313 |
| KF294371 | China | 2013-01-01 | 0.6192 | Rattus losea | -356040368.5036 |
| MH687971 | Viet Nam | 2014-11-12 | 0.6194 | Rattus argentiventer | -356161570.0562 |
| MH687977 | Viet Nam | 2015-02-04 | 0.6199 | Rattus argentiventer | -356466591.1490 |
| KF294357 | China | 2011-01-01 | 0.6214 | Apodemus agrarius | -357298941.1683 |
| KM349744 | China | 2012-05-17 | 0.6219 | Rattus norvegicus (Norway rat) | -357570433.8433 |
| NC_026011 | China | 2012-05-17 | 0.6219 | Rattus norvegicus (Norway rat) | -357570433.8433 |
| KM349743 | China | 2012-05-17 | 0.6220 | Rattus norvegicus (Norway rat) | -357646895.7536 |

\* qdistance: Smaller values implies higher risk
† Likelihood lower bound: Larger values implies higher risk
‡ RaTG13

**TABLE XVI.** Numbering Conversion to pdm09 and H3 Schemes

| Query | H1N1pdm | H3 | Query | H1N1pdm | H3 | Query | H1N1pdm | H3 | Query | H1N1pdm | H3 |
| --- | --- | --- | --- | --- | --- | --- | --- | --- | --- | --- | --- |
| 1 | - | - | 77 | 60 | 69 | 157 | 140 | 143 | - | - | - |
| 2 | - | - | 78 | 61 | 70 | 158 | 141 | 144 | - | - | - |
| 3 | - | - | 79 | 62 | 71 | 159 | 142 | 145 | - | - | - |
| 4 | - | - | 80 | 63 | 72 | 160 | 143 | 146 | 238 | 221 | 224 |
| 5 | - | - | 81 | 64 | 73 | 161 | 144 | 147 | 239 | 222 | 225 |
| 6 | - | - | 82 | 65 | 74 | 162 | 145 | 148 | 240 | 223 | 226 |
| 7 | - | - | 83 | 66 | 75 | 163 | 146 | 149 | 241 | 224 | 227 |
| 8 | - | - | 84 | 67 | 76 | 164 | 147 | 150 | 242 | 225 | 228 |
| 9 | - | - | 85 | 68 | 77 | 165 | 148 | 151 | 243 | 226 | 229 |
| 10 | - | - | 86 | 69 | 78 | 166 | 149 | 152 | 244 | 227 | 230 |
| 11 | - | - | 87 | 70 | 79 | 167 | 150 | 153 | 245 | 228 | 231 |
| 12 | - | - | 88 | 71 | 80 | 168 | 151 | 154 | 246 | 229 | 232 |
| 13 | - | - | 89 | 72 | 81 | 169 | 152 | 155 | 247 | 230 | 233 |
| 14 | - | - | 90 | 73 | 82 | 170 | 153 | 156 | 248 | 231 | 234 |
| 15 | - | - | 91 | 74 | - | 171 | 154 | 157 | 249 | 232 | 235 |
| 16 | - | - | 92 | 75 | 83 | 172 | 155 | 158 | 250 | 233 | 236 |
| 17 | - | - | 93 | 76 | 84 | - | - | - | 251 | 234 | 237 |
| - | - | 1 | 94 | 77 | 85 | - | - | - | 252 | 235 | 238 |
| - | - | 2 | 95 | 78 | 86 | - | - | - | 253 | 236 | 239 |
| - | - | 3 | 96 | 79 | 87 | - | - | - | 254 | 237 | 240 |
| - | - | 4 | 97 | 80 | 88 | 173 | 156 | 159 | 255 | 238 | 241 |
| - | - | 5 | 98 | 81 | 89 | 174 | 157 | 160 | 256 | 239 | 242 |
| - | - | 6 | 99 | 82 | 90 | 175 | 158 | 161 | 257 | 240 | 243 |
| - | - | 7 | 100 | 83 | 91 | 176 | 159 | 162 | 258 | 241 | 244 |
| - | - | 8 | 101 | 84 | 92 | 177 | 160 | 163 | 259 | 242 | 245 |
| - | - | 9 | 102 | 85 | - | 178 | 161 | 164 | 260 | 243 | 246 |
| - | - | 10 | 103 | 86 | 93 | 179 | 162 | 165 | 261 | 244 | 247 |
| 18 | 1 | 11 | 104 | 87 | 94 | 180 | 163 | 166 | 262 | 245 | 248 |
| 19 | 2 | 12 | 105 | 88 | 95 | 181 | 164 | 167 | 263 | 246 | 249 |
| 20 | 3 | 13 | 106 | 89 | 96 | 182 | 165 | 168 | 264 | 247 | 250 |
| 21 | 4 | 14 | 107 | 90 | 97 | 183 | 166 | 169 | 265 | 248 | 251 |
| 22 | 5 | 15 | 108 | 91 | 98 | 184 | 167 | 170 | 266 | 249 | 252 |
| 23 | 6 | 16 | 109 | 92 | 99 | - | - | - | 267 | 250 | 253 |
| 24 | 7 | 17 | 110 | 93 | 100 | 185 | 168 | 171 | 268 | 251 | 254 |
| 25 | 8 | 18 | 111 | 94 | 101 | 186 | 169 | 172 | 269 | 252 | 255 |
| 26 | 9 | 19 | 112 | 95 | 102 | 187 | 170 | 173 | 270 | 253 | 256 |
| 27 | 10 | 20 | - | - | - | - | - | - | 271 | 254 | 257 |
| 28 | 11 | 21 | - | - | - | 188 | 171 | 174 | 272 | 255 | 258 |
| 29 | 12 | 22 | 113 | 96 | 103 | 189 | 172 | 175 | 273 | 256 | 259 |
| 30 | 13 | 23 | 114 | 97 | 104 | 190 | 173 | 176 | 274 | 257 | 260 |
| 31 | 14 | 24 | 115 | 98 | 105 | 191 | 174 | 177 | 275 | 258 | 261 |
| 32 | 15 | 25 | 116 | 99 | 106 | 192 | 175 | 178 | 276 | 259 | 262 |
| 33 | 16 | 26 | 117 | 100 | 107 | 193 | 176 | 179 | - | - | - |
| 34 | 17 | 27 | 118 | 101 | 108 | 194 | 177 | 180 | - | - | - |
| 35 | 18 | 28 | 119 | 102 | 109 | 195 | 178 | 181 | - | - | - |
| 36 | 19 | 29 | 120 | 103 | 110 | 196 | 179 | 182 | - | - | - |
| 37 | 20 | 30 | 121 | 104 | 111 | 197 | 180 | 183 | - | - | - |
| 38 | 21 | 31 | 122 | 105 | 112 | 198 | 181 | 184 | - | - | - |
| 39 | 22 | 32 | 123 | 106 | 113 | 199 | 182 | 185 | - | - | - |
| 40 | 23 | 33 | 124 | 107 | 114 | 200 | 183 | 186 | - | - | - |
| 41 | 24 | 34 | 125 | 108 | 115 | 201 | 184 | 187 | - | - | - |
| 42 | 25 | 35 | 126 | 109 | 116 | 202 | 185 | 188 | 277 | 260 | - |
| 43 | 26 | 36 | 127 | 110 | 117 | 203 | 186 | 189 | 278 | 261 | 263 |
| 44 | 27 | 37 | 128 | 111 | 118 | 204 | 187 | 190 | 279 | 262 | 264 |
| 45 | 28 | 38 | 129 | 112 | 119 | 205 | 188 | 191 | 280 | 263 | 265 |
| 46 | 29 | 39 | 130 | 113 | 120 | 206 | 189 | 192 | 281 | 264 | 266 |
| 47 | 30 | 40 | 131 | 114 | 121 | 207 | 190 | 193 | 282 | 265 | 267 |
| 48 | 31 | 41 | 132 | 115 | 122 | 208 | 191 | 194 | 283 | 266 | 268 |
| 49 | 32 | 42 | 133 | 116 | 123 | 209 | 192 | 195 | 284 | 267 | 269 |
| 50 | 33 | 43 | - | - | - | 210 | 193 | 196 | 285 | 268 | 270 |
| 51 | 34 | 44 | - | - | - | 211 | 194 | 197 | 286 | 269 | 271 |
| 52 | 35 | 45 | 134 | 117 | 124 | 212 | 195 | 198 | 287 | 270 | 272 |
| 53 | 36 | 46 | 135 | 118 | 125 | 213 | 196 | 199 | 288 | 271 | 273 |
| 54 | 37 | 47 | 136 | 119 | - | - | - | - | 289 | 272 | 274 |
| 55 | 38 | 48 | 137 | 120 | - | 214 | 197 | 200 | 290 | 273 | 275 |
| 56 | 39 | 49 | 138 | 121 | - | 215 | 198 | 201 | 291 | 274 | 276 |
| 57 | 40 | 50 | 139 | 122 | 126 | 216 | 199 | 202 | 292 | 275 | 277 |
| 58 | 41 | 51 | 140 | 123 | 127 | 217 | 200 | 203 | 293 | 276 | 278 |
| 59 | 42 | 52 | 141 | 124 | 128 | 218 | 201 | 204 | 294 | 277 | 279 |
| 60 | 43 | 53 | - | - | - | 219 | 202 | 205 | 295 | 278 | 280 |
| 61 | 44 | 54 | - | - | - | 220 | 203 | 206 | 296 | 279 | 281 |
| 62 | 45 | - | - | - | - | 221 | 204 | 207 | 297 | 280 | 282 |
| 63 | 46 | 55 | - | - | - | 222 | 205 | 208 | 298 | 281 | 283 |
| 64 | 47 | 56 | - | - | - | 223 | 206 | 209 | 299 | 282 | 284 |
| 65 | 48 | 57 | 142 | 125 | 129 | 224 | 207 | 210 | 300 | 283 | 285 |
| 66 | 49 | 58 | 143 | 126 | 130 | 225 | 208 | 211 | - | - | - |
| 67 | 50 | 59 | 144 | 127 | 131 | 226 | 209 | 212 | 301 | 284 | 286 |
| 68 | 51 | 60 | 145 | 128 | 132 | 227 | 210 | 213 | 302 | 285 | 287 |
| - | - | - | 146 | 129 | 133 | 228 | 211 | 214 | 303 | 286 | 288 |
| - | - | - | 147 | 130 | - | 229 | 212 | 215 | 304 | 287 | 289 |
| - | - | - | 148 | 131 | 134 | 230 | 213 | 216 | 305 | 288 | 290 |
| - | - | - | 149 | 132 | 135 | 231 | 214 | 217 | 306 | 289 | 291 |
| - | - | - | 150 | 133 | 136 | 232 | 215 | 218 | 307 | 290 | 292 |
| 69 | 52 | 61 | 151 | 134 | 137 | 233 | 216 | 219 | 308 | 291 | 293 |
| 70 | 53 | 62 | 152 | 135 | 138 | 234 | 217 | 220 | 309 | 292 | 294 |
| 71 | 54 | 63 | 153 | 136 | 139 | 235 | 218 | 221 | 310 | 293 | 295 |
| 72 | 55 | 64 | 154 | 137 | 140 | 236 | 219 | 222 | 311 | 294 | 296 |
| 73 | 56 | 65 | 155 | 138 | 141 | 237 | 220 | 223 | - | - | - |
| 74 | 57 | 66 | - | - | - | - | - | - | 312 | 295 | 297 |
| 75 | 58 | 67 | 156 | 139 | 142 | - | - | - | 313 | 296 | 298 |

## References

[1] Fair, J. & Fair, J. Viral forecasting, pathogen cataloging, and disease ecosystem mapping: Measuring returns on investments (2019).

[2] Hannenhalli, S. & Pevzner, P. Transforming cabbage into turnip.(polynomial algorithm for sorting signed permutations by reversals). dept. of computer science and engineering, penn state university. Tech. Rep., Technical Report CSE-95-004 (1995).

[3] Jean, G. & Nikolski, M. Genome rearrangements: a correct algorithm for optimal capping. Information Processing Letters 104, 14–20 (2007).

[4] Ozery-Flato, M. & Shamir, R. Two notes on genome rearrangement. Journal of Bioinformatics and Computational Biology 1, 71–94 (2003).

[5] Tesler, G. Efficient algorithms for multichromosomal genome rearrangements. Journal of Computer and System Sciences 65, 587–609 (2002).

[6] Shao, M. & Lin, Y. Approximating the edit distance for genomes with duplicate genes under dcj, insertion and deletion. BMC bioinformatics 13, S13 (2012).

[7] Rulli, M. C., Santini, M., Hayman, D. T. & D’Odorico, P. The nexus between forest fragmentation in africa and ebola virus disease outbreaks. Scientific reports 7, 41613 (2017).

[8] Chua, K. B., Chua, B. H. & Wang, C. W. Anthropogenic deforestation, el niiio and the emergence of nipah virus in malaysia. Malaysian Journal of Pathology 24, 15–21 (2002).

[9] Childs, J. Zoonotic viruses of wildlife: hither from yon. In Emergence and Control of Zoonotic Viral Encephalitides, 1–11 (Springer, 2004).

[10] Gamblin, S. J. & Skehel, J. J. Influenza hemagglutinin and neuraminidase membrane glycoproteins. Journal of Biological Chemistry 285, 28403–28409 (2010).

[11] Bosch, B. J., van der Zee, R., de Haan, C. A. & Rottier, P. J. The coronavirus spike protein is a class i virus fusion protein: structural and functional characterization of the fusion core complex. Journal of virology 77, 8801–8811 (2003).

[12] Cheverud, J. M. & Routman, E. J. Epistasis and its contribution to genetic variance components. Genetics 139, 1455–1461 (1995).

[13] Cleaveland, S., Laurenson, M. K. & Taylor, L. H. Diseases of humans and their domestic mammals: pathogen characteristics, host range and the risk of emergence. Philos. Trans. R. Soc. Lond., B, Biol. Sci. 356, 991–999 (2001).

[14] Wolfe, N. D., Daszak, P., Kilpatrick, A. M. & Burke, D. S. Bushmeat hunting, deforestation, and prediction of zoonoses emergence. Emerging Infect. Dis. 11, 1822–1827 (2005).

[15] Holmes, E. C. & Drummond, A. J. The evolutionary genetics of viral emergence. Curr. Top. Microbiol. Immunol. 315, 51–66 (2007).

[16] Parrish, C. R. et al. Cross-species virus transmission and the emergence of new epidemic diseases. Microbiol. Mol. Biol. Rev. 72, 457–470 (2008).

[17] Childs, J. E. & Gordon, E. R. Surveillance and control of zoonotic agents prior to disease detection in humans. Mt. Sinai J. Med. 76, 421–428 (2009).

[18] Pulliam, J. R. & Dushoff, J. Ability to replicate in the cytoplasm predicts zoonotic transmission of livestock viruses. J. Infect. Dis. 199, 565–568 (2009).

[19] Pepin, K. M., Lass, S., Pulliam, J. R., Read, A. F. & Lloyd-Smith, J. O. Identifying genetic markers of adaptation for surveillance of viral host jumps. Nat. Rev. Microbiol. 8, 802–813 (2010).

[20] Flanagan, M. L. et al. Anticipating the Species Jump: Surveillance for Emerging Viral Threats. Zoonoses and Public Health 59, 155–163 (2012). 15334406.

[21] Cover, T. M. & Thomas, J. A. Elements of Information Theory (Wiley-Interscience, New York, NY, USA, 1991).

[22] Varadhan, S. S. Large deviations. In Proceedings of the International Congress of Mathematicians 2010 (ICM 2010) (In 4 Volumes) Vol. I: Plenary Lectures and Ceremonies Vols. II–IV: Invited Lectures, 22–639 (World Scientific, 2010).

[23] White, J. M., Delos, S. E., Brecher, M. & Schornberg, K. Structures and mechanisms of viral membrane fusion proteins: multiple variations on a common theme. Critical reviews in biochemistry and molecular biology 43, 189–219 (2008).

[24] Boni, M. F. Vaccination and antigenic drift in influenza. Vaccine 26, C8–C14 (2008).

[25] Agor, J. K. & Ö zaltin, O. Y. Models for predicting the evolution of influenza to inform vaccine strain selection. Human vaccines & immunotherapeutics 14, 678–683 (2018).

[26] (2020). URL https://www.cdc.gov/flu/vaccines-work/effectiveness-studies.htm.

[27] van der Meer, F. J. U. M., Orsel, K. & Barkema, H. W. The new influenza A H1N1 virus: balancing on the interface of humans and animals. The Canadian veterinary journal = La revue veterinaire canadienne 51, 56–62 (2010).

[28] Smith, G. J. D. et al. Origins and evolutionary genomics of the 2009 swine-origin H1N1 influenza A epidemic. Nature 459, 1122–1125 (2009).

[29] Zhou, P. et al. A pneumonia outbreak associated with a new coronavirus of probable bat origin. Nature 579, 270–273 (2020).

[30] Andersen, K. G., Rambaut, A., Lipkin, W. I., Holmes, E. C. & Garry, R. F. The proximal origin of sars-cov-2. Nature medicine 26, 450–452 (2020).

[31] Carugo, O. & Pongor, S. A normalized root-mean-spuare distance for comparing protein three-dimensional structures. Protein science 10, 1470–1473 (2001).

[32] Righetto, I., Milani, A., Cattoli, G. & Filippini, F. Comparative structural analysis of haemagglutinin proteins from type a influenza viruses: conserved and variable features. BMC bioinformatics 15, 363 (2014).

[33] Lee, B. & Richards, F. M. The interpretation of protein structures: estimation of static accessibility. Journal of molecular biology 55, 379–IN4 (1971).

[34] Shrake, A. & Rupley, J. A. Environment and exposure to solvent of protein atoms. lysozyme and insulin. Journal of molecular biology 79, 351–371 (1973).

[35] Momen-Roknabadi, A., Sadeghi, M., Pezeshk, H. & Marashi, S.-A. Impact of residue accessible surface area on the prediction of protein secondary structures. Bmc Bioinformatics 9, 357 (2008).

[36] Adamczak, R., Porollo, A. & Meller, J. Combining prediction of secondary structure and solvent accessibility in proteins. Proteins: Structure, Function, and Bioinformatics 59, 467–475 (2005).

[37] Tzarum, N. et al. Structure and receptor binding of the hemagglutinin from a human h6n1 influenza virus. Cell host & microbe 17, 369–376 (2015).

[38] Lazniewski, M., Dawson, W. K., Szczepin’ska, T. & Plewczynski, D. The structural variability of the influenza a hemagglutinin receptor-binding site. Briefings in functional genomics 17, 415–427 (2018).

[39] Garcia, N. K., Guttman, M., Ebner, J. L. & Lee, K. K. Dynamic changes during acid-induced activation of influenza hemagglutinin. Structure 23, 665–676 (2015).

[40] Cleveland, W. S. Lowess: A program for smoothing scatterplots by robust locally weighted regression. American Statistician 35, 54 (1981).

## References

[1] Bosch, B. J., van der Zee, R., de Haan, C. A. & Rottier, P. J. The coronavirus spike protein is a class i virus fusion protein: structural and functional characterization of the fusion core complex. Journal of virology 77, 8801–8811 (2003).

[2] McAuley, J., Gilbertson, B., Trifkovic, S., Brown, L. E. & McKimm-Breschkin, J. Influenza virus neuraminidase structure and functions. Frontiers in microbiology 10, 39 (2019).

[3] Hatcher, E. L. et al. Virus variation resource–improved response to emergent viral outbreaks. Nucleic acids research 45, D482–D490 (2017).

[4] Bogner, P., Capua, I., Lipman, D. J. & Cox, N. J. A global initiative on sharing avian flu data. Nature 442, 981–981 (2006).

[5] Herna’ndez-Orozco, S., Kiani, N. A. & Zenil, H. Algorithmically probable mutations reproduce aspects of evolution, such as convergence rate, genetic memory and modularity. Royal Society open science 5, 180399 (2018).

[6] Cover, T. M. & Thomas, J. A. Elements of Information Theory (Wiley-Interscience, New York, NY, USA, 1991).

[7] Hothorn, T., Hornik, K. & Zeileis, A. Unbiased recursive partitioning: A conditional inference framework. JOURNAL OF COMPUTATIONAL AND GRAPHICAL STATISTICS 15, 651–674 (2006).

[8] Manning, C. D., Manning, C. D. & Sch ü tze, H. Foundations of statistical natural language processing (MIT press, 1999).

[9] Fedotov, A. A., Harremo ë s, P. & Topsoe, F. Refinements of pinsker’s inequality. IEEE Transactions on Information Theory 49, 1491–1498 (2003).

